# Sample Size Requirements for Machine Learning Classification of Binary Outcomes in Bulk RNA-Seq Data

**DOI:** 10.1101/2025.08.19.25333999

**Authors:** Scott Silvey, Amy Olex, Shaojun Tang, Jinze Liu

**Affiliations:** Virginia Commonwealth University, School of Public Health, Department of Biostatistics. Richmond VA 23219; Virginia Commonwealth University, Wright Center for Clinical and Translational Research. Richmond VA 23219

## Abstract

Bulk RNA sequencing data is often leveraged to build machine learning (ML)-based predictive models for classification of disease groups or subtypes, but the sample size needed to adequately train these models is unknown. We collected 27 experimental datasets from the Gene Expression Omnibus and the Cancer Genome Atlas. In 24/27 datasets, pseudo-data were simulated using Bayesian Network Generation. Three ML algorithms were assessed: XGBoost (XGB), Random Forest (RF), and Neural Networks (NN). Learning curves were fit, and sample sizes needed to reach the full-dataset AUC minus 0.02 were determined and compared across the datasets/algorithms. Multivariable negative binomial regression models quantified relationships between dataset-level characteristics and required sample sizes within each algorithm. These models were validated in independent experimental datasets. Across the datasets studied, median required sample sizes were 480 (XGB)/190 (RF)/269 (NN). Higher effect sizes, less class imbalance/dispersion, and less complex data were associated with lower required sample size. Validation demonstrated that predictions were accurate in new data. Comparison of results to sample sizes obtained from differential analysis power analysis methods showed that ML methods generally required larger sample sizes. In conclusion, incorporating ML-based sample size planning alongside traditional power analysis can provide more robust results.

## Introduction

High-dimensional data contains significantly more measured variables than samples. -Omics data, a type of high-dimensional data, is generated from objective physiological processes or markers typically measured in serum, plasma, or other sample types [1]. One popular domain within -omics is bulk RNA sequencing (RNA-Seq), which quantifies gene expression levels by measuring the abundance of messenger RNA [2]. Over the past two decades, there have been significant advances in the technology used to collect this type of data, leading to a rise in publications aiming to link specific gene signatures to a clinical condition or outcome of interest [3]. This is often achieved through the use of ML algorithms, which are becoming increasingly popular in clinical decision making due to their promising gain in performance over traditional linear models [4]. For example, Yu et. al (2020) used ML to build an 8-gene classifier for pancreatic cancer, showing that their derived gene panel was superior to CA19-9 (gold standard) in discriminating early-stage disease from healthy controls or chronic pancreatitis [5].

RNA-Seq data differs from clinical or electronic health record-based data in several ways. First, predictors are numerous and may be highly correlated due to intrinsic properties of the genes themselves [6]. Also, the features generated from RNA-Seq data are count-based in nature and right-skewed due to the lack of an upper boundary of gene expression [7]. Most importantly, RNA-Seq data is expensive to collect, with a single sample costing potentially hundreds of dollars to run [8]. Because of this, RNA-Seq studies in humans typically contain limited sample sizes compared to clinical data, which is often gathered from a database or other record system, does not usually require additional costs to accrue for researchers, and is typically recorded for primary purposes other than research [9].

Studies aiming to develop predictive models using RNA-Seq data can follow many potential pipelines, but a general approach involves a two-step process - after data cleaning and pre-processing has been completed - beginning with a differential analysis through the use of formal statistical testing, regularization, or a supervised non-parametric approach. This aims to reduce the feature space to only genes that are statistically significant or clinically relevant, often referred to as a gene signature [10]. Once a set of differentially expressed genes is established, machine learning models are trained using these features. This allows for a parsimonious model that can be more easily applied to a clinical setting [11].

Within the scope of RNA-Seq data, the statistical power needed to perform parametric differential analysis through hypothesis testing is well-studied, with multiple tools available for calculation of the necessary sample size needed to detect a pre-set number of significant genes given assumptions regarding the effect sizes, variation, proportion of differentially expressed genes, and other aspects of the data [12]. In contrast, the literature is sparse regarding how much *additional* sample, if any, is needed in order to train a machine learning model that provides stable predictions. While the sample size needed for identifying individual differentially expressed genes may be low, training ML models may require more observations due to the multivariable nature of these analyses, as well as interactive and other nonlinear effects between genes/biomarkers and the clinical outcome or endpoint.

Understanding how sample size requirements differ for the use of ML methods and standard differential analysis approaches is fundamental for study design, future reproducibility, and cost-effective use of limited data. Thus, we aim to perform a novel examination of sample size-requirements across three popular machine learning algorithms and a variety of experimental RNA-Seq datasets. We provide an in-depth exploration of dataset-level characteristics and how they influence required sample sizes, eventually creating and validating a tool that can be used to predict the required sample size in a new dataset.

## Results

### Dataset Description and Algorithm Performance

We gathered 24 Gene Expression Omnibus (GEO) datasets with original sample sizes ranging from 32 to 474 (Table 1), differential analysis techniques were used to filter out important genes, and then Bayesian network generation was used to simulate large artificial data using these sources – the specific approach is detailed in the Methods and Supplementary Section A. Figure 1 details the entire methodological process of our study. The GEO datasets spanned 9 clinical subspecialties and disease categories including cardiovascular, pulmonary, autoimmune, cancer, and others (Table 1). From the Cancer Genome Atlas (TCGA), we utilized 3 datasets without simulating pseudo-data, of which specific information can be found in the Methods section. The binary clinical outcomes examined within all 27 datasets can be seen in Table 1.

**Figure 1.**
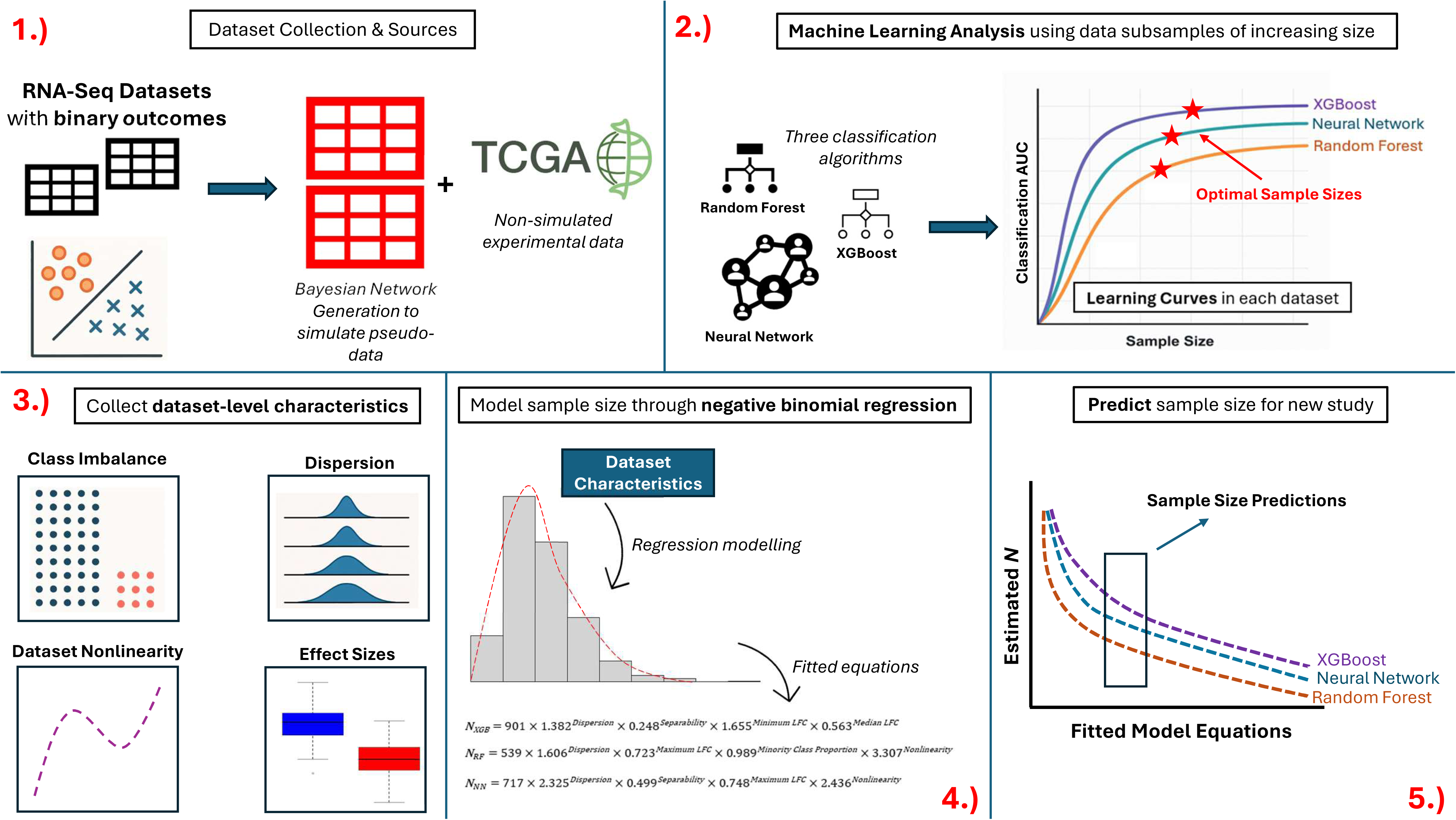
**Methodological Process and Study Overview** Flowchart of methodology and utility of final methods discussed in this manuscript. Panel 1.) Describes the data collection and simulation procedure used to generate large pseudo-data. Panel 2.) Shows learning curve creation and classification algorithms Panel 3.) Details dataset-level characteristics to be used in model building. Panel 4.) Shows the model fitting process. Panel 5.) Shows how models can be applied to new datasets for sample size determination.

**Table 1.** Dataset Overview.

| *Dataset Name* | *Size* | *GEO Accession/TCGA ID* | *Comparison (Condition vs. Reference)* | *Disease Category* |
| --- | --- | --- | --- | --- |
| GBM | 32 | GSE188812 | Long vs. Short Survival in High-Grade Glioma | Cancer |
| HCC | 63 | GSE114564 | Early vs. Late-Stage Hepatocellular Carcinoma | Cancer |
| Kidney | 82 | GSE218048 | IF/TA post-Kidney Transplant vs. Healthy | Kidney |
| IPF | 84 | GSE124685 | Idiopathic Pulmonary Fibrosis vs. Healthy | Pulmonary |
| COVID | 84 | GSE184610 | COVID-19 vs. Healthy | Pulmonary |
| HBV | 95 | GSE173897 | Hmong vs. Other Ethnicities in HBV-related HCC | Cancer |
| Hypertension | 97 | GSE183635 | Angina Pectoris vs. Pulmonary Hypertension | Cardiovascular |
| NAFLD | 98 | GSE167523 | NASH vs. NAFL | Liver |
| PrePost | 99 | GSE229083 | Pre vs. Post-capillary Pulmonary Hypertension | Pulmonary |
| RA | 100 | GSE117769 | Rheumatoid Arthritis vs. Healthy | Autoimmune |
| AVSC | 101 | GSE218474 | Aortic Valve Sclerosis vs. no-AVSC | Cardiovascular |
| Crohn | 125 | GSE137344 | Inflammatory Bowel Disease vs. Healthy | Gastro |
| MDD | 139 | GSE80655 | Major Depression vs. Healthy | Psychiatric |
| Ovarian | 148 | GSE230522 | Aspirin Use vs. No-Aspirin in Ovarian Cancer | Cancer |
| ALS | 162 | GSE124439 | ALS vs. Healthy | Neurological |
| Kawasaki | 165 | GSE178491 | Kawasaki Disease vs. Febrile Control | Autoimmune |
| CCA | 170 | GSE183635 | Cholangiocarcinoma vs. Colorectal Cancer | Cancer |
| Glioma | 176 | GSE183635 | Glioma vs. Epilepsy | Cancer |
| TBProg | 177 | GSE103147 | Tuberculosis Disease Progression vs. No Progression | Pulmonary |
| Tuberculosis | 182 | GSE89403 | Tuberculous Cure vs. No-Cure | Pulmonary |
| EDS | 200 | GSE218012 | Hypermobility Disease vs. Healthy | Autoimmune |
| NSCLC | 218 | GSE207586 | Non-Small Cell Lung Cancer vs. Healthy | Cancer |
| Bipolar | 239 | GSE124326 | Lithium vs. No-Lithium in Bipolar Disorder | Psychiatric |
| MS | 474 | GSE183635 | Multiple Sclerosis vs. Healthy | Autoimmune |
| TCGA-Brain | 875 | TCGA-GBM, TCGA-LGG | Glioblastoma vs. Low Grade Glioma | Cancer |
| TCGA-Breast | 925 | TCGA-BRCA | Invasive Ductal Carcinoma vs. Other Pathology | Cancer |
| TCGA-Lung | 1162 | TCGA-LUAD, TCGA-LUSC | Lung Adenocarcinoma vs. Lung Squamous Cell Carcinoma | Cancer |

Of the three classification algorithms examined, Neural Networks (NN) performed the best or tied for the best area-under the receiver operating characteristic curve performance (AUC) on 23/27 (85.2%) of the datasets, while XGBoost performed the best on 3 (“HCC,” “PrePost,” and “TCGA-Lung”), and Random Forest was only superior in the TCGA-Breast dataset.

AUCs calculated using the entire datasets ranged from 0.718-1.000 (Table 2). Throughout the manuscript, the terms “full-dataset AUC”, “maximum achievable AUC”, and “separability” were used interchangeably. 11 of the 27 datasets (40.7%) had full-dataset AUCs > 0.99, which indicated nearly perfect separability as the sample size became very large. It is important to note that these AUCs were calculated under the assumption that the validation sets were drawn from the same population as the training sets, which may explain the reasoning behind these high values [13]. If researchers aim to apply ML models to a validation set derived from a different population than the training set, larger sample sizes may be necessary [14].

**Table 2.**
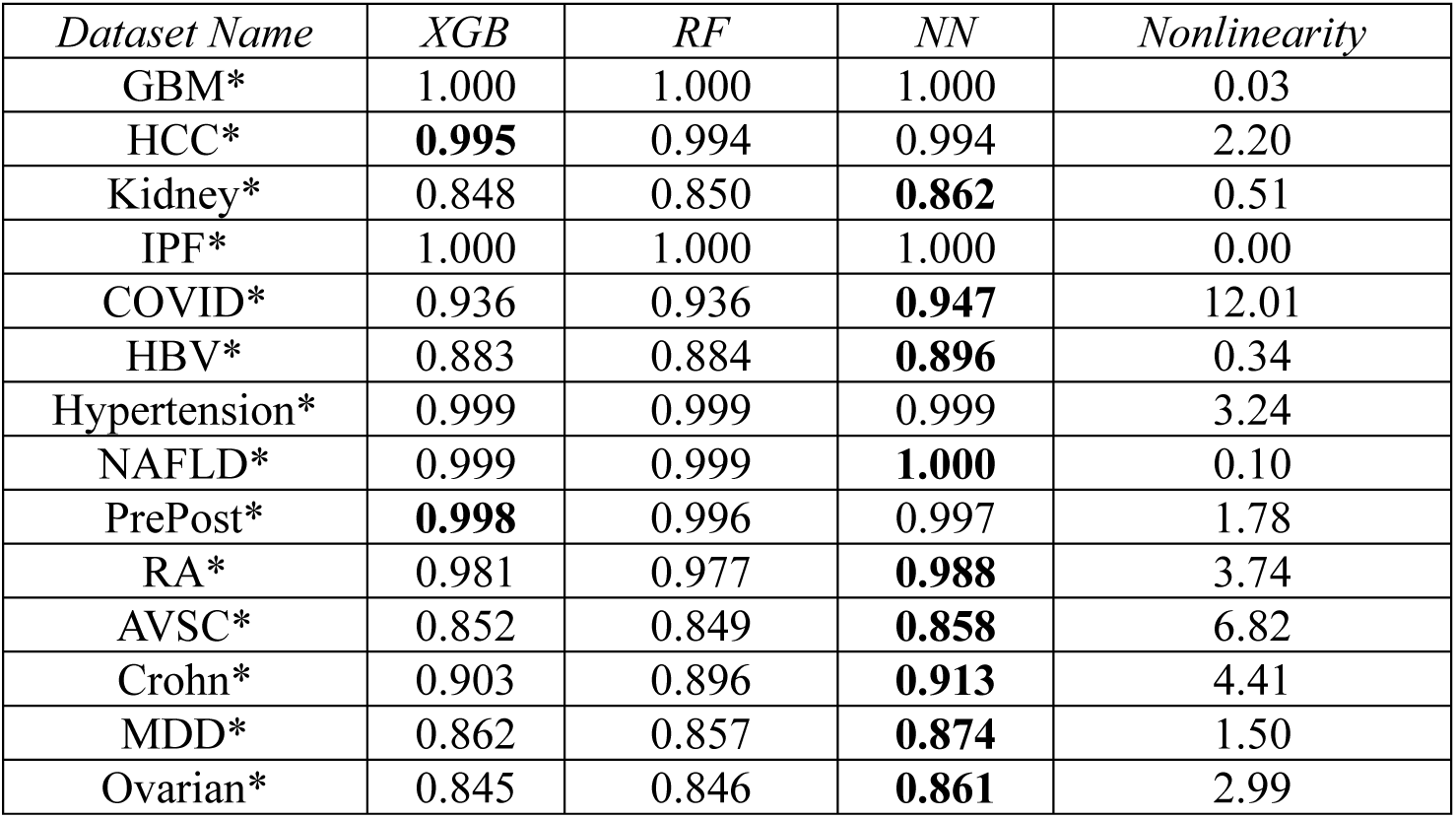
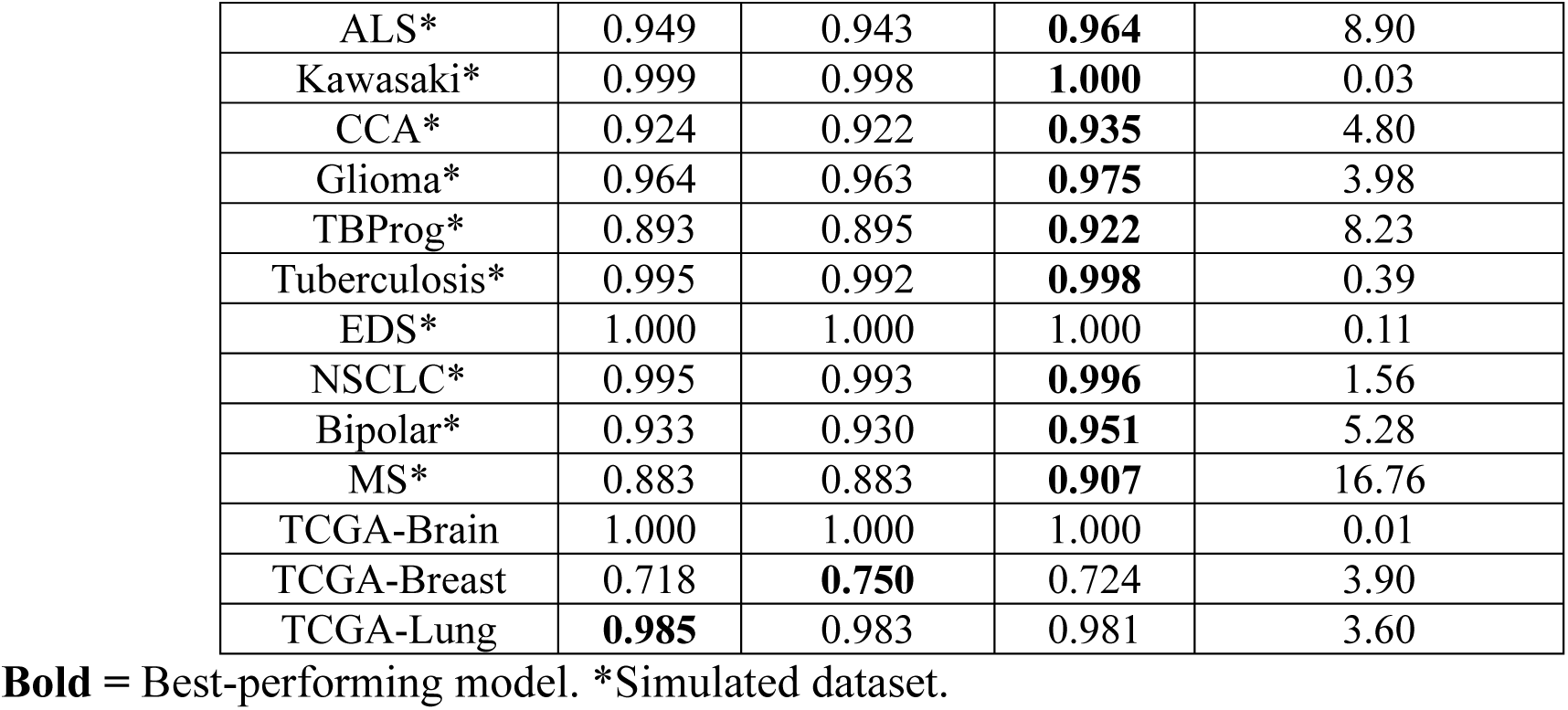
Full Dataset AUCs over Three ML Algorithms.

### Sample Size Requirements Vary by Algorithm

Figure 2 shows required sample sizes for the three algorithms over all datasets, and a table of specific values can be found in supplementary section B. XGB required the largest sample sizes (median *n* = 480, range: 54-1,200), while RF required the least (median *n* = 190, range: 25-1,307). Neural Network sample sizes were the most variable (median *n* = 269, range: 25-1,711).

**Figure 2.**
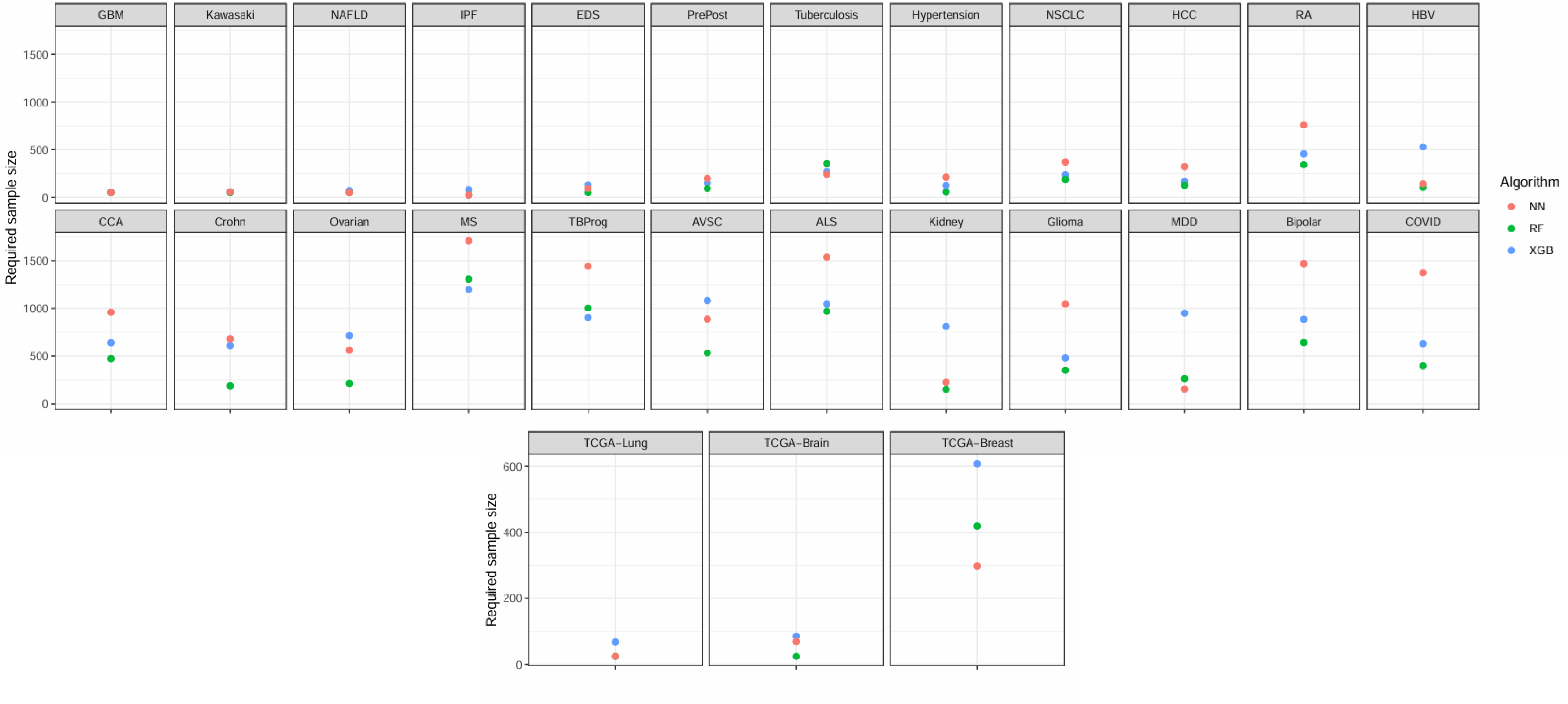
**Sample Size Visualization** Displays the required sample sizes needed to obtain AUC within 0.02 of maximum achievable value within all 27 datasets used for model fitting. The top panel contains the datasets which were simulated, and the bottom panel contains the three TCGA datasets which were used directly without simulation.

### Summary of Dataset-Level Characteristics Across Experimental Studies

Dataset-level characteristics were collected (Supplementary Section B, Table 3). The average minority class percentage was 34.94% (SD: ± 13.66%) and the average number of differentially expressed genes included in ML modelling was 24 (± 29). Among the differentially expressed genes, average median log-fold change (LFC) was 1.36 (± 1.08), minimum LFC was 0.48 (± 0.58), and maximum LFC was 3.45 (± 2.50). The average median dispersion among the differentially expressed genes was 1.47 (± 2.50), the average read count was 650.88 (± 1669.33), and average between-gene correlation was 0.37 (± 0.19).

**Table 3.** Dataset-Level Characteristics.

| *Dataset Name* | *Minority Class Proportion* | *Number of Differentially Expressed Genes* | *Median LFC\* (Min, Max)* | *Median Between-Gene Correlation\** | *Median Dispersion\** | *Median Average Read Count\** |
| --- | --- | --- | --- | --- | --- | --- |
| GBM | 43.8% | 8 | 1.12 (0.23, 2.13) | 0.36 | 0.32 | 1220.70 |
| HCC | 28.6% | 9 | 2.32 (0.88, 5.64) | 0.55 | 1.33 | 252.10 |
| Kidney | 50.0% | 3 | 0.43 (0.32, 1.44) | 0.13 | 0.88 | 83.39 |
| IPF | 41.7% | 58 | 2.96 (0.74, 7.63) | 0.67 | 0.03 | 276.40 |
| COVID | 43.5% | 5 | 3.71 (3.17, 4.92) | 0.54 | 9.65 | 8772.62 |
| HBV | 30.5% | 3 | 0.86 (0.56, 2.32) | 0.08 | 0.14 | 51.22 |
| Hypertension | 26.8% | 13 | 1.64 (0.42, 4.70) | 0.57 | 1.31 | 63.21 |
| NAFLD | 48.0% | 52 | 0.59 (0.27, 2.32) | 0.54 | 0.12 | 384.34 |
| PrePost | 49.5% | 16 | 1.10 (0.39, 2.61) | 0.32 | 0.42 | 56.50 |
| RA | 49.0% | 12 | 0.34 (0.21, 1.06) | 0.44 | 0.08 | 334.02 |
| AVSC | 39.6% | 3 | 1.15 (0.92, 2.23) | 0.33 | 2.05 | 31.14 |
| Crohn | 17.6% | 4 | 1.09 (0.19, 2.00) | 0.06 | 0.55 | 30.34 |
| MDD | 49.6% | 4 | 0.45 (0.33, 0.66) | 0.46 | 0.16 | 144.81 |
| Ovarian | 39.9% | 5 | 0.60 (0.34, 2.24) | 0.22 | 0.35 | 380.08 |
| ALS | 10.5% | 8 | 0.73 (0.15, 2.50) | 0.14 | 2.87 | 17.29 |
| Kawasaki | 17.6% | 41 | 1.48 (0.16, 5.33) | 0.63 | 0.36 | 316.41 |
| CCA | 50.0% | 9 | 1.52 (0.46, 2.99) | 0.24 | 1.51 | 1041.63 |
| Glioma | 25.0% | 11 | 1.37 (0.52, 2.84) | 0.26 | 1.01 | 82.72 |
| TBProg | 17.5% | 7 | 1.19 (0.57, 3.45) | 0.17 | 9.74 | 118.69 |
| Tuberculosis | 7.7% | 13 | 0.42 (0.20, 1.43) | 0.26 | 0.07 | 1342.51 |
| EDS | 50.0% | 71 | 0.86 (0.20, 5.16) | 0.32 | 0.39 | 314.77 |
| NSCLC | 23.4% | 37 | 1.25 (0.67, 2.76) | 0.43 | 0.23 | 276.82 |
| Bipolar | 36.4% | 16 | 0.64 (0.21, 1.95) | 0.66 | 0.07 | 112.21 |
| MS | 18.1% | 12 | 0.39 (0.09, 1.18) | 0.33 | 0.80 | 90.04 |
| TCGA-Brain | 41.0% | 102 | 3.39 (0.43, 8.62) | 0.60 | 1.49 | 35.49 |
| TCGA-Breast | 39.6% | 18 | 0.83 (0.18, 1.90) | 0.25 | 0.84 | 615.84 |
| TCGA-Lung | 48.5% | 103 | 4.42 (0.21, 11.19) | 0.56 | 2.86 | 1128.37 |

\*Among those which were considered to be truly differentially expressed.

Dataset nonlinearity was calculated by taking the difference in AUC between the best-performing ML method and a separately fitted multivariable logistic regression model (LR) using the same feature set. Median dataset nonlinearity was 2.74 (± 3.60), indicating that ML approaches performed 2.74 AUC-points better than LR, on average. For future modelling, due to the difficulty of correctly guessing this value without already knowing properties of the full dataset, we transformed nonlinearity into a categorical variable, with a threshold of 4.5, allowing for only two potential options for downstream sample size estimation. This threshold minimized the out-of-sample mean squared error, and a detailed description of how this was obtained can be found in supplementary section F. About one-quarter of the datasets (7/27, 25.9%) had nonlinearity values over the determined high threshold of 4.5 AUC points.

Other dataset characteristics that were categorized for the purpose of downstream modelling included the maximum achievable AUC (<0.99 vs. ≥0.99), median between-gene correlation (<0.5 vs. ≥0.5), and median dispersion (<1 vs. ≥1). These characteristics were categorized in order to simplify the final models and allow for less exhaustive estimation of sample sizes if specific pilot data is not available.

### Dataset-Level Characteristics are Strongly Associated with Sample Sizes

Negative binomial regression models were fit, examining the individual associations between each of the dataset-level characteristics and predicted sample sizes. For XGBoost, separability, number of differentially expressed genes, between-gene correlation, median LFC, and maximum LFC were inversely related with sample size, while nonlinearity was positively associated with increased sample size. For datasets with high separability (max./full-dataset AUC > 99.0), estimated sample sizes were affected by a multiplier of 0.177 (*p*<0.001). For every additional differentially expressed gene, estimated sample sizes were affected by a multiplier of 0.975 (*p*<0.001). For a one-unit increase in maximum LFC, estimated sample sizes were affected by a multiplier of 0.766 (*p*<0.001). For a one-unit increase in median LFC, estimated sample sizes were affected by a multiplier of 0.698 (*p*=0.014). In datasets with high (≥ 4.5) values of nonlinearity, estimated sample sizes were affected by a multiplier of 2.741 (*p*=0.002). Finally, for datasets with average between-gene correlation greater than 0.5, estimated sample sizes were affected by a multiplier of 0.400 (*p*=0.005). In the Neural Network analyses, results were similar for separability (0.180× multiplier for high, *p*<0.001), nonlinearity (4.793× multiplier for high, *p*<0.001), and number of differentially expressed genes (0.968× multiplier for every additional predictor, *p*<0.001). For Random Forest, minority class proportion (0.973× multiplier for every 1% increase, *p*=0.044), separability (0.211× multiplier for high, *p*<0.001), nonlinearity (4.850× multiplier for high, *p*<0.001), number of differentially expressed genes (0.968× multiplier for every additional predictor, *p*<0.001), correlation (0.397× multiplier for median correlation > 0.5, *p*=0.016), median LFC (0.666× multiplier for a 1-unit increase, *p*=0.018), and max. LFC (0.716× multiplier for every 1-unit increase, *p*<0.001) were significantly associated with sample size with effect directions identical to the other two algorithms. Figure 3 visualizes some of these relationships, and the remainder of the plots can be found in Supplementary Section G. In multivariable models for each algorithm, we selected the set of up to four predictors that minimized Akaike’s Information Criterion (AIC). Equation 1 (below) shows model estimates.

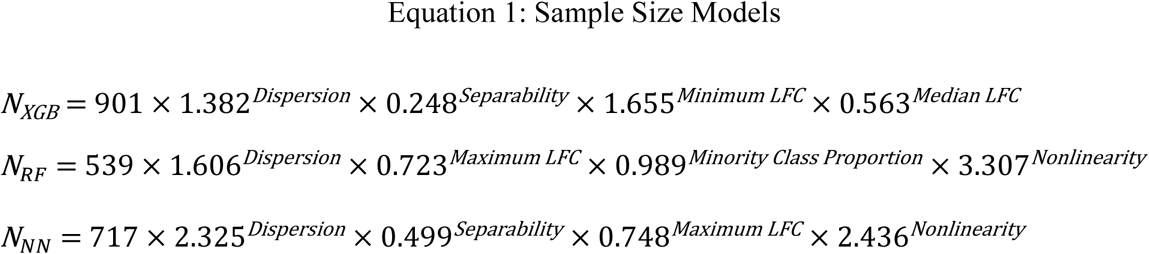

**Figure 3.**
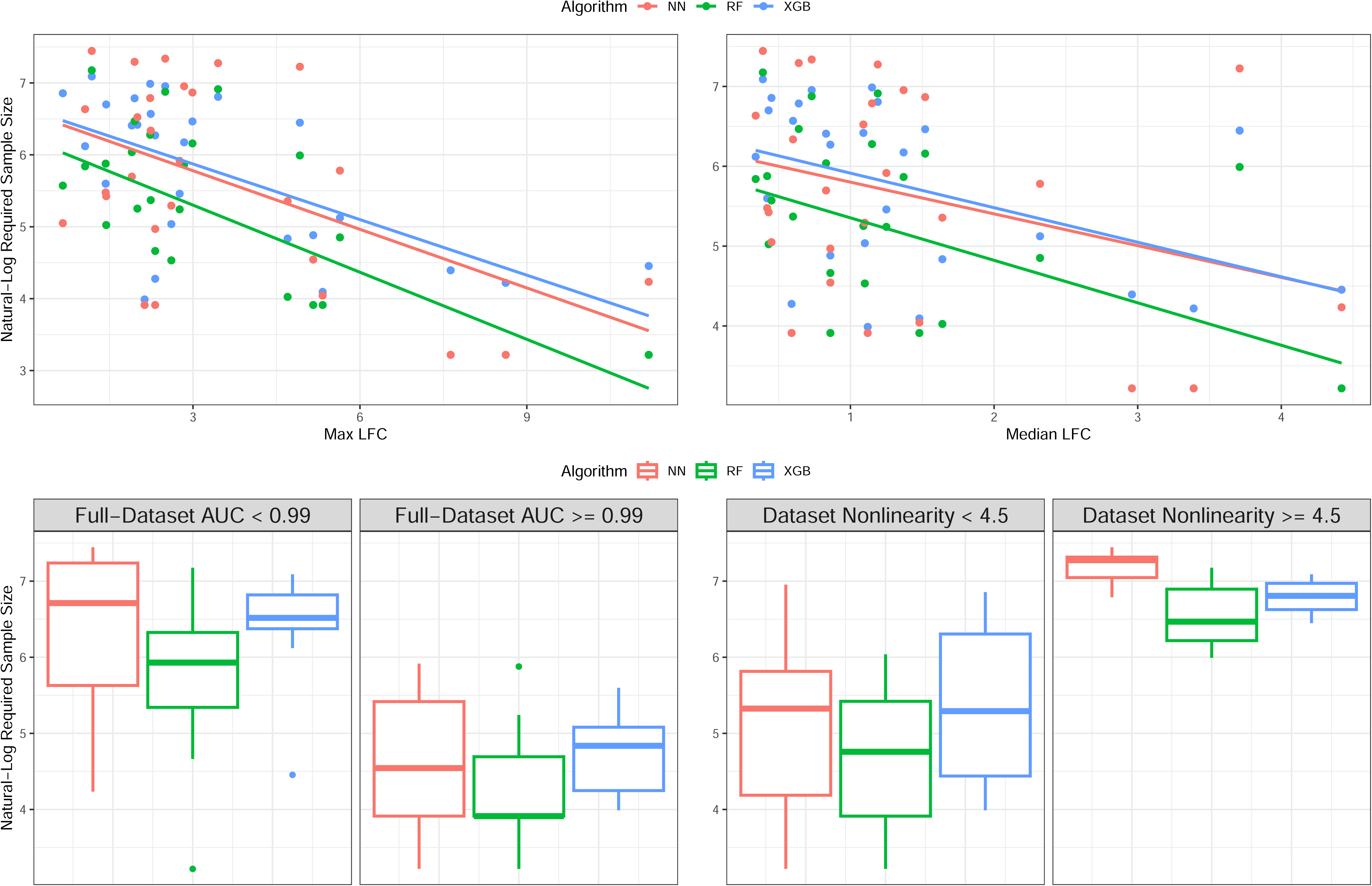
**Univariable Relationships Between Dataset-Level Characteristics and Sample Size** Displays univariable relationships between four impactful dataset-level variables : separability (max. achievable AUC), Max Log-fold change, Median log-fold change, and dataset nonlinearity. Y-axis is the sample size transformed by natural-log. Each color represents a different machine learning algorithm.

Supplementary section G contains tables showing univariable and multivariable contributions of each predictor to the expected sample size, along with *p*-value and 95% confidence intervals. Figure 4 visualizes fitted model estimates within each final multivariable model.

**Figure 4.**
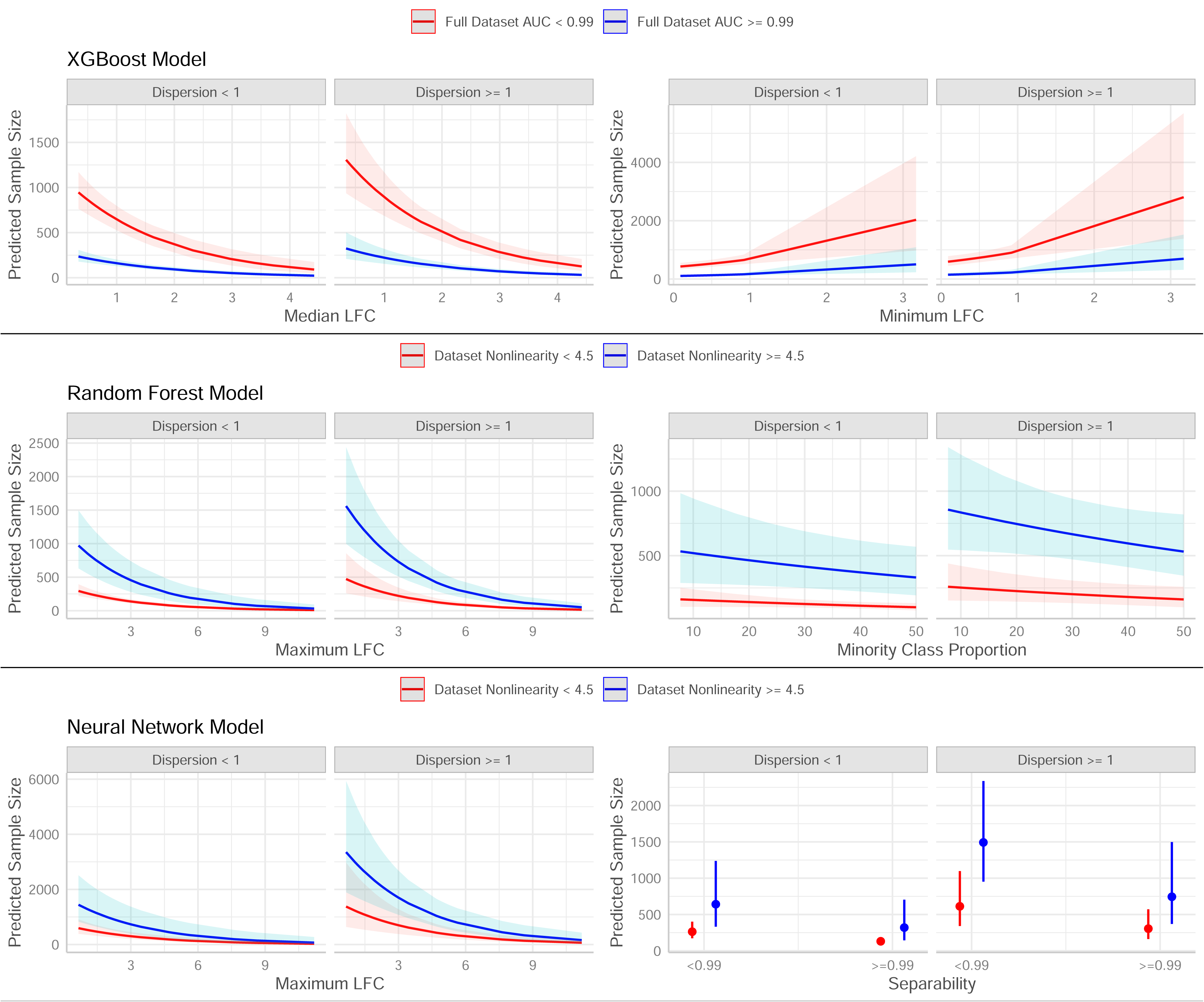
**Multivariable Model Estimates** Displays fitted model estimates and 95% confidence intervals generated by multivariable sample size models. Each model used four dataset-level predictors, so effects are shown for varying levels of each characteristic. Y-axis is the sample size prediction.

For all three models, median dispersion was selected as a top predictor. For RF and NN models, dataset nonlinearity was a top predictor. For XGB and NN, dataset separability was a top predictor. Other included variables were minimum LFC and median LFC (XGB model), and max. LFC (RF/NN models). For those which were also statistically significant from univariable analysis, the direction and magnitude of multivariable coefficient estimates was similar (supplementary section G). Deviance-based R^2^ statistics, adjusted for the number of predictors added, were 0.841 (XGB), 0.793 (RF), and 0.687 (NN), indicating that the dataset-level predictors explained a majority of the total deviance.

Among the 27 datasets, 10 (37.0%) were cancer related (Table 1). In order to test the equivalence of model effects among cancer and non-cancer datasets, we fit predictor-by-disease state interaction effects and assessed statistical significance. We found no evidence of significant interaction effects between dataset-characteristics and cancer vs. non-cancer outcomes in all three ML-specific models.

### Sample Size Prediction Models can be Reliably Applied to New Experimental Datasets

Two independent GEO datasets were collected – which we called “PDAC” and “IBD” [5,15], containing 339 and 2471 observations respectively. Dataset characteristics from these two validation sets can be found in Table 4.

**Table 4.** Validation Dataset Overview.

| Summary: |  |  |  |  |  |
| --- | --- | --- | --- | --- | --- |
| Dataset Name | GEO Accession | Sample Size (Full, Train/Test) | Comparison | Disease Subtype |  |
| IBD | GSE193677 | 2471 (2000/471) | Irritable Bowel Disease (UC/Crohn's) vs. Healthy | Gastro |  |
| PDAC | GSE133684 | 339 (300/39) | Pancreatic Cancer vs. Healthy | Cancer |  |
| Characteristics*: |  |  |  |  |  |
| Dataset Name | Minority Class Proportion | Full-Dataset AUC | Median LFC (Min, Max) | Median Dispersion | Nonlinearity |
| IBD | 18.1% | **XGB: 0.917, RF: 0.911, NN: 0.929** | 0.90 (0.17, 2.62) | 1.07 | Low |
| PDAC | 30.1% | **XGB: 1.000, RF: 1.000, NN: 1.000** | 0.87 (0.01, 5.58) | 0.89 | Low |

\*Only those needed to calculate $n$ from sample size models are listed. Others can be found in
Supplementary Section D.

When our sample size models were applied using these dataset-characteristics as input, we predicted required *n* for XGB/RF/NN of 809 (95% CI: [597-1,096]), 301 (95% CI: [178-511]), and 779 (95% CI: [402-1,513]) in “IBD,” respectively, compared to true required sample sizes of 836, 367, and 905 calculated directly from learning curves. In “PDAC,” we predicted required n for XGB/RF/NN of 136 (95% CI: [107-173]), 63 (95% CI: [43-91]), and 71 (95% CI: [44-113]) respectively, compared to true required sample sizes of 176, 80, and 113. Learning curves can be seen in Figure 5. In five out of six predicted scenarios, the true sample size was included in the 95% CI of the predicted *n*, and the average difference between true and predicted sample size was ±53 observations. This represented the ability of our models to predict the correct sample size within a narrow range. A full summary of the validation results can be found in Table 5. As a sensitivity analysis, our sample size models were re-fit by swapping IBD and PDAC for the three TCGA datasets, and assessing model performance on these TCGA sets. We found that model estimates and validation accuracy obtained from the re-fit model were similar to those presented in the main text (Supplementary Section C).

**Figure 5.**
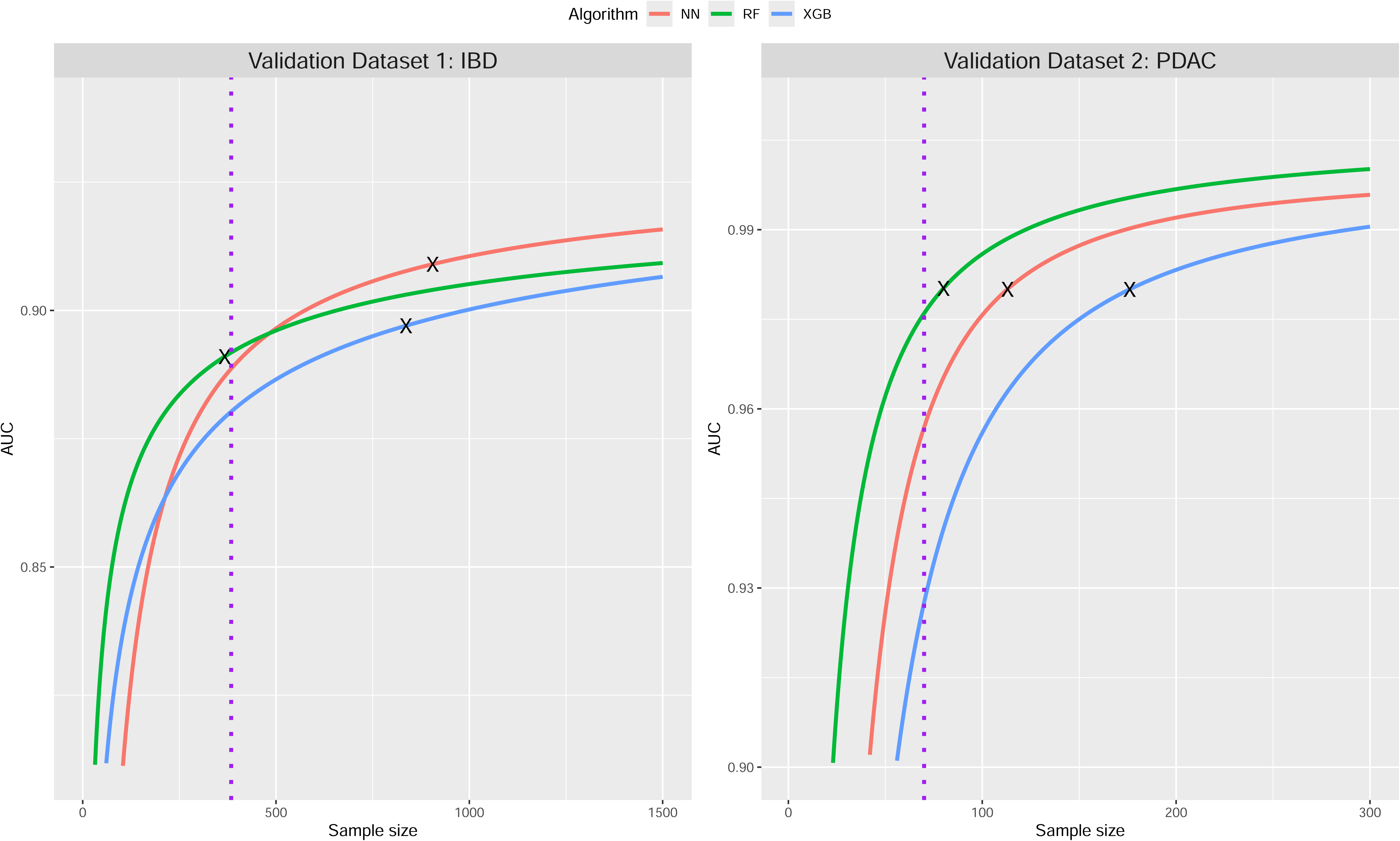
**Learning Curves for Validation Datasets** Figure Caption – Fitted learning curves for RF (green), XGB (blue), and NN (red) algorithms in the two validation datasets. Black X indicates the sample size where AUC stability was reached using our definition of max. achievable AUC – 0.02. Purple dotted line is the sample size required to achieve 80% power in differential expression calculated using the ssizeRNA package.

**Table 5.**
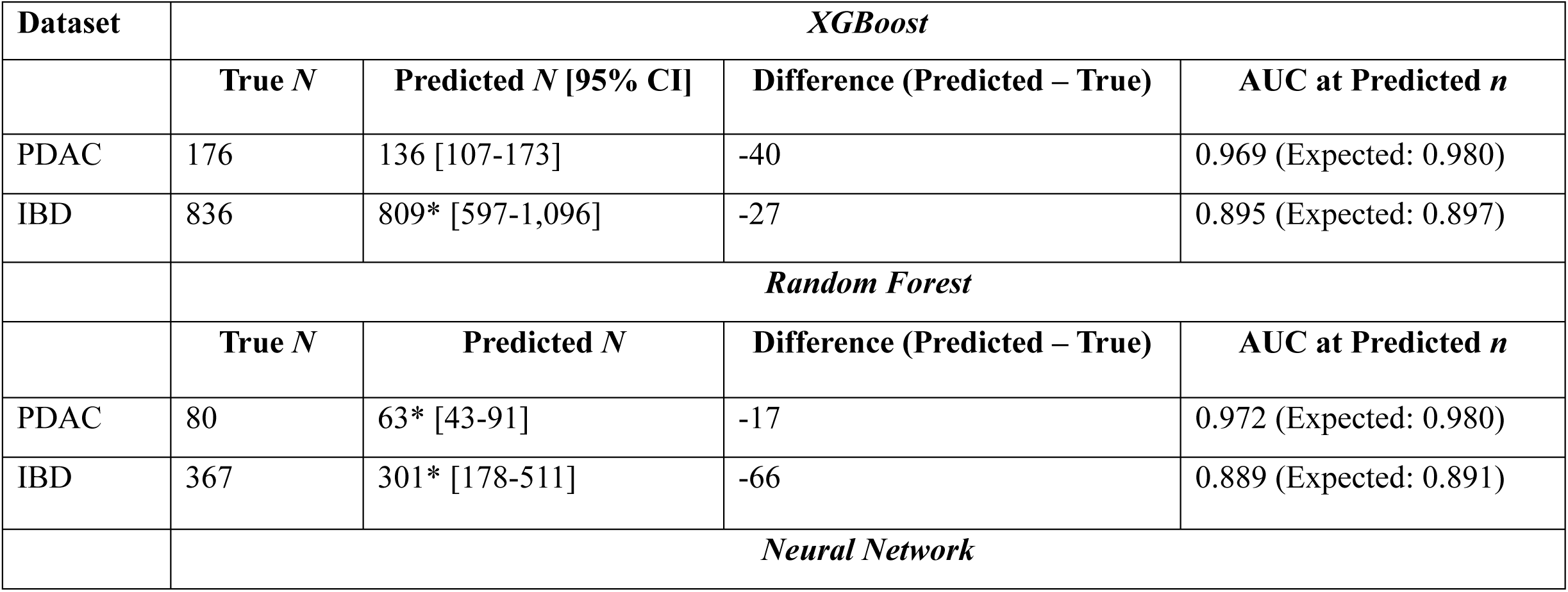
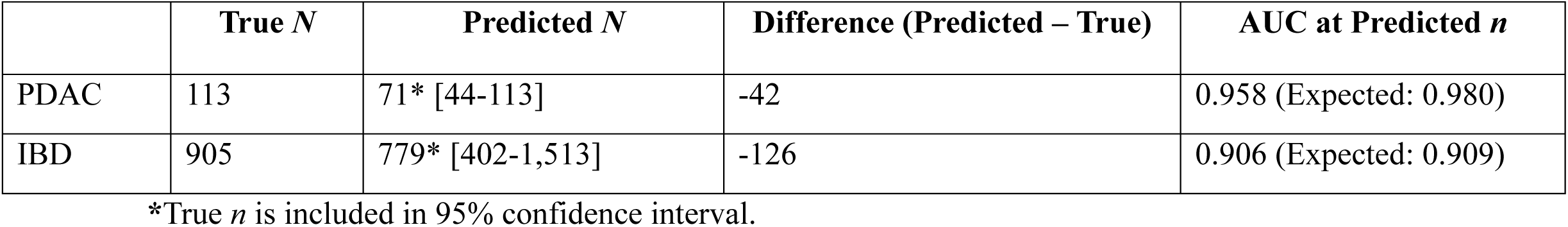
Validation Results.

### Machine Learning Requires More Observations than Traditional Power Analysis

We have previously established that the sample size required for differential analysis is well-studied [12]. Using the *ssizeRNA* R package [16], we calculated the sample sizes required to detect the differentially expressed genes used in ML modelling of our validation datasets, “IBD” and “PDAC,” and compared these sample sizes to the *n* required for machine learning. We found that “IBD” required approximately 384 samples to attain 80% power to detect these genes, while “PDAC” required approximately 70 samples. These numbers were slightly higher than the *n* required for Random Forest in both datasets (Table 5/Figure 5, IBD: 367, PDAC: 80). At an *n* of 384 and using a neural network model - the best-performing algorithm within “IBD” in the unrestricted dataset - an AUC of 0.889 was achieved, which represents a 4.0-point decrease from the maximum achievable value (0.929), but Random Forest was able to perform better at this smaller sample size, achieving an AUC of 0.891. In “PDAC,” where the max. achievable AUC value was 1 over all three tested methods, an *n* of 70 was sufficient to achieve an AUC of 0.975 using a Random Forest model (2.5-point decrease from max.) but only 0.956/0.957 with NN/XGB. Depending on budgetary concerns, it is possible that a slightly lower AUC versus the maximum achievable value may be acceptable in order to prioritize costs. Additionally, these results demonstrated that RF tended to provide stronger AUC results at lower sample sizes but is eventually outperformed by NN/XGB in larger samples. While NN was generally the superior predictive modelling approach among the datasets collected in this study at an unrestricted sample size, a Random Forest model may be optimal if data collection is heavily restricted by costs.

## Discussion

In this study, we performed a learning curve analysis of 27 RNA-Seq datasets (24 simulated, 3 non-simulated) over three different ML algorithms. From this, we identified the expected samples needed to consistently obtain AUCs that were within two points of the maximum achievable values. We then examined the effects of dataset-level characteristics on these sample sizes and provided models based on these characteristics that can be used to predict the necessary sample size in a new dataset.

We found that random forest consistently required the smallest sample sizes (median n = 190), XGBoost (median n = 480) required the largest sample sizes, and neural networks required a sample size that was between RF and XGB but had the largest variability (median n = 269). Neural networks also consistently provided the highest maximum achievable AUCs in the full datasets among the three methods tested. Interestingly, for certain datasets (see: TCGA-Breast, MDD, etc.), NN was the algorithm that required the least sample size. This phenomenon highlights the variability in output over all three ML algorithms.

Dataset-level variables that altered expected sample sizes varied by algorithm, but the overall separability of the dataset, the effect sizes and variability of the features, and nonlinearity were among the most influential characteristics. In validation datasets, we demonstrated that our models were able to reliably estimate required sample sizes. Although further validation is needed, this demonstrated the promise of our method to accurately predict required *n* in varying cases. This also demonstrated the validity of our simulation approach, since the models derived using mostly pseudo-data were able to perform well on non-simulated RNA-Seq data.

Most of the dataset-level characteristics examined can be reasonably guessed or obtained before an experiment begins. For example, researchers can examine prior studies in their field of interest or use already-collected pilot data to determine a reasonable range for minority class proportion, log-fold changes of key predictors, and dispersion.

For separability, obtaining this value requires knowing properties of the entire dataset, which is not feasible in experimental scenarios. However, one can examine prior studies comparing similar outcomes, or use intuition regarding the clinical comparison of interest itself - comparisons that are more subtle may have lower maximum performance compared to more obvious comparisons. For example, in our datasets, GSE173897 (comparison of ethnic groups in HBV-originated hepatocellular carcinoma) had a full-dataset AUC of only 0.896, whereas in GSE207586 (comparison of non-small cell lung cancer vs. healthy patients [17]), we obtained an AUC of 0.996. Datasets with high separability also had more differentially expressed genes identified, with average feature set sizes of 14 (low-separability) vs. 38 (high-separability).

Finally, if pilot data is available, ML performance can be assessed at a limited sample size. For example, in our datasets, we found that those which ultimately ended up having high separability (or maximum achievable AUC > 0.99) were all able to attain a training-set AUC > 0.90 at *n* = 25.

Dataset nonlinearity is less intuitive to guess prior to data collection. In general, we found that datasets with nonlinearity values of at least 4.5 (meaning that, the best ML algorithm outperforms logistic regression by at least 4.5 AUC points) required approximately 2-3 times larger sample sizes, depending on algorithm. In our datasets, nonlinearity is highly linked to separability. Specifically, datasets with a max. AUC ≥ 0.99 were all low-nonlinearity. This is intuitive, because once the feature set is extremely strong, the AUC will be maxed out, regardless of interactions, dependencies, etc. However, in datasets with max. AUC < 0.99, around half (9/16, 56%) of the datasets had low nonlinearity values. Once more, prior studies where both simple (LR) and complex (NN/RF/XGB) methods are compared can help researchers determine if this value will be high. As a last resort, since we modelled dataset nonlinearity as a categorical variable, researchers can simply calculate expected sample sizes for both scenarios (high vs. low nonlinearity) using our model equations presented in this study and select the scenario that is most cost-effective.

To our knowledge, no prior study has presented specific formulas for calculating sample size within the context of machine learning for RNA-Seq experiments. Current applications of ML to RNA-Seq data are not explicitly powered for the use of these models, and thus may be collecting too few samples, sacrificing potential performance gain. Comparison of our results to a differential expression power analysis method showed that XGBoost and Neural Network required larger sample sizes, while Random Forest did not require many additional observations beyond those needed for standard differential analysis. Additionally, Random Forest had the strongest AUC when restricted to the differential analysis sample size, but collecting more data produced higher max. achievable AUCs among NN and XGB algorithms. We believe that all of these results should be taken into consideration when designing future RNA-Seq experiments. Specifically, if researchers are interested in using ML approaches in an -omics setting, they should calculate sample sizes needed for differential expression *and* machine-learning and use both estimates, as well as potential budgetary limitations, to inform data collection.

## Limitations and Future Work

One limitation of our study was the inability to generalize these results to an external validation scenario, i.e. application of ML models to unseen data drawn from a separate population compared to the training data. We believe that if external validation is considered for an experiment, then the required sample size will be higher, but the exact amounts are unclear. This is one potential avenue for future research. Another limitation was reliance on simulated data– using only non-simulated data would provide more clarity and confidence in the final results. However, prior studies that examined sample size requirements for RNA-Seq differential analysis also relied heavily on data simulation [16,18], since the amount of large open-source experimental datasets available is limited.

Future research in this area could examine other -omics disciplines such as metabolomics or proteomics. These additional methods are becoming more prevalent, due to their ability to complement RNA-Seq data by allowing for analysis of downstream protein or metabolic products affected by the genes of interest [19]. The steps needed to replicate this experiment and fit new sample size models for other -omics disciplines could be easily implemented by adjusting the simulation procedure to more closely resemble the data structure of interest, as long as adequate datasets are available. Additionally, extension to different clinical outcomes such as continuous, multi-class, or survival endpoints would be impactful.

## Methods

### Dataset Description

Experimental RNA-Seq datasets were collected from two public sources - the Gene Expression Omnibus (GEO) and the Cancer Genome Altas (TCGA) [20,21]. GEO is a repository containing gene expression data submitted by researchers in a common format that is publicly accessible to query and download. TCGA is another public repository containing gene expression information and metadata from over 20,000 human samples and 33 cancer types. Both repositories have been widely utilized to discover novel biomarkers and classify disease subtypes [22,23].

We collected 26 GEO datasets spanning a variety of clinical domains such as pulmonary disease, cardiovascular disease, mental health, and cancer (Table 1). We collected three datasets from TCGA which included lung cancer, brain cancer, and breast cancer – these were the three largest disease domains available [21]. The lung cancer dataset (which we define as “TCGA-Lung”) contained observations pooled from two sources, TCGA-LUSC and TCGA-LUAD, two lung cancer subtypes [24]. The brain cancer dataset (which we define as “TCGA-Brain”) contained pooled samples from TCGA-LGG (low-grade glioma) and TCGA-GBM [25,26] (glioblastoma). Finally, the breast cancer dataset contained only data from the TCGA-BRCA study [23]. The clinical comparison examined within TGCA-Lung was the LUAD vs. LUSC subtypes. For TCGA-Brain, we compared low-grade glioma vs. the more aggressive glioblastoma. Finally, the clinical comparison within TCGA-BRCA was invasive ductal carcinoma (IDC) vs. all other subtypes. Of note, we were not able to obtain information related to triple-negative breast cancer status, which is a known comparison of interest due to differences in treatability between these classes [27].

### Learning Curves are used to Measure Performance at Increase Sample Sizes

It is known that, for any given dataset, there is a maximum achievable predictive capability that can be attained, regardless of the number of observations included [28]. This concept has been explored extensively and is achieved by creating a learning curve, which measures a performance metric as a function of sample size [29]. Learning curves allow for an examination of a model’s predictive ability at increasing training set sizes, and subsequent identification of a point of diminishing returns where data collection can reasonably be stopped.

The choice of predictive metric used when creating learning curves is subjective. However, one fair and widely used performance metric is the area under the receiver operating characteristic curve (AUC), which evaluates predicted probabilities over a range of thresholds from 0 to 1 [30]. AUC is used to evaluate the discriminative performance of an ML algorithm in a new sample and is insensitive to the proportion of cases versus controls in the dataset [31].

### Data Simulation is Used to Generate Pseudo-Data

In order to build accurate learning curves, it is necessary to fully understand the behavior of the AUC as *n* approaches a very large size. However, access to large open-source RNA-Seq datasets is limited – for example, most GEO datasets contain <100 total samples. We addressed this by using Bayesian Network Generation (BNG) to generate pseudo-data. BNG is a statistical learning technique that aims to model a set of variables and their conditional dependencies using Directed Acyclic Graphs (DAG) and has been shown to produce data that is structurally similar to the original source [32]. Full details can be found in supplementary section A.

Within each original dataset and prior to simulation, we determined a set of differentially expressed genes, defined as all genes in the original source that were both 1.) statistically significant through the DESeq2 pipeline [33] using FDR-adjusted *p*<0.05, and 2.) included when the Boruta [34] algorithm was applied. DESeq2 was selected over other pipelines because it has been shown to provide the best performance when the number of samples is greater than 6 [PMID: 36112652]. Boruta is a nonparametric variable selection algorithm that returns a feature set that is unbiased and contains all important attributes within the dataset, according to authors [34].

All pseudo-data had n=5,000 observations with outcome distributions equal to those in the original sources. Only the differentially expressed gene sets were simulated. Additional data pre-processing steps included removal of features with zero-variance and DESeq2 normalization [33]. Consistency of the simulation approach in recovering the proper effect sizes and accurate data structure was examined visually (Supplementary Section E). 24/26 GEO datasets required this simulation approach, and all three TCGA datasets were large enough to be studied without simulation.

Prior studies examining sample size requirements for RNA-Seq differential analysis have also employed simulation, typically using a negative binomial distribution with pre-specified parameters [16,18]. However, we chose to use BNG to simulate pseudo-data because this approach is able to capture interactions or unknown relationships between genes, which is not possible with a strict negative binomial simulation approach.

### Overview of Machine Learning Methods Used

We examined the following ML algorithms: Random Forest (RF) [35], XGBoost (XGB) [36], and Neural Networks (NN) [37]. These methods were selected due to their widespread and popular use in both -omics and clinical data analysis [38]. For RF and XGB, hyperparameters were left at their default values, which can be found in the R documentation [35,36]. For neural networks, we used the R package h2o [39] to perform our analyses; we considered one hidden layer with 100 units, and 10 epochs of data training. We chose these hyperparameters in order to study a fixed configuration of a basic neural network architecture - but discuss the potential implications of changing hyperparameters. The type of neural network used by “h2o” is a multi-layer feedforward artificial neural network, also known as multilayer perceptron (MLP). The activation function used in the hidden layers was the default linear rectifier, with SoftMax activation in the final output nodes for probability estimation and classification. Prior to NN analysis, we centered and scaled all predictors to ensure optimal performance. Other neural network hyperparameters were again left at their default values, which can be found in the documentation [39].

### Defining Required Sample Sizes from Learning Curves

In each dataset, we evaluated the mean out-of-sample AUC as a function of increasing sample size. The specific learning curve approach is detailed in Supplementary Section B. In summary, we took a random sample of size *n* from the full dataset, trained each ML model on this subsample, and then tested model performance on another independent sample. At each size *n*, this process was repeated 50 times, and the test-set AUC was averaged to obtain a final estimate. Then, we increased *n* and repeated the process. Finally, using standard approaches (see: Supplementary section B), smooth curves were fit to the raw data points, which we refer to as learning curves.

Our stopping rule was focused on achieving AUC stability, or a plateauing of the curve which indicated future data collection would not increase performance greatly. We defined this as the smallest *n* where the mean out-of-sample AUC was within two points (0.02) of the observed full-dataset (or, maximum achievable) AUC. For example, if the maximum achievable AUC was 0.95, we would obtain the smallest *n* where the mean out-of-sample AUC of 0.93 was first surpassed. The maximum achievable AUC for each classification algorithm was estimated using five-fold outcome-stratified cross-validation (CV). We chose a stopping point of 0.02 because we believed that it optimized the trade-off between performance and sample size based on prior research [40]. However, once the curves are generated, any user-selected threshold can be applied. To demonstrate this, we present a supplementary analysis using alternative thresholds of one-point (0.01) and five-points (0.05).

### Overview of Dataset-Level Characteristics

Following the learning curve analysis of the three ML algorithms, we measured the effects of 10 dataset-level factors on the sample sizes needed for AUC stability. A full description of these factors and their definitions can be found in Supplementary Section B. In summary, we examined minority class proportion, separability (defined as the full-dataset AUC), the total number of differentially expressed genes, median/maximum/minimum log-fold change (LFC) among the differentially expressed genes, median between-gene correlation, median dispersion, median average read count, and dataset nonlinearity. Dataset nonlinearity was an estimate of the degree of nonlinear or interactive relationships between the predictors and the outcome that were present in the data. This was defined as the point difference in the maximum-achievable AUC when using the best-performing ML algorithm compared to a multivariable logistic regression model [40]. For example, if logistic regression yielded an AUC of 0.90 and XGBoost yielded an AUC of 0.95, the dataset nonlinearity would be calculated as 5. A higher value of nonlinearity indicates a more complex dataset, with regards to the relationships between the predictors and the response variable. Once more, for the purpose of calculating these values, hyperparameters were left at their default values for all ML methods.

### Creating Sample Size Prediction Models

Within the context of each ML algorithm, the relationship between dataset-level variables and the required sample sizes was examined. Because the estimated sample sizes were discrete and right-skewed, we used negative binomial regression models [41] to quantify the strength and significance of each dataset characteristic on expected sample size, which produce coefficients in terms of log-expected counts. Then, in multivariable models for each algorithm, we selected the combination of predictors which together minimized the model’s Akaike Information Criterion (AIC), which quantifies goodness-of-fit while penalizing for additional covariates [42].

We also calculated adjusted deviance-based pseudo-*R*^2^ statistics [43,44], which further quantified each model’s goodness-of-fit and proportion of deviance explained by the predictors. The equivalence of model coefficients was tested between cancer vs. non-cancer datasets through tests of interaction, since there are known differences in expression levels and structure of cancer-related RNA-Seq data [45].

The final model equations were reported and discussed for each algorithm, and visualizations of predictions at varying levels of the fitted model equations were generated.

Statistical significance was set to α = 0.05 for all hypothesis tests considered, and RStudio version 4.4.1 was used for all analyses.

### Validation of Sample Size Models on New Experimental Data

As noted previously, 24/26 GEO datasets were artificially expanded through simulation. In our main analysis, we used these 24 simulated datasets and all three original TCGA datasets to fit our sample size models. This left two remaining non-simulated GEO datasets for model validation.

The first dataset was GSE193677 (IBD) which compared 2,471 patients with either Crohn’s disease, ulcerative colitis, or neither [15]. The second dataset was GSE133684 (PDAC) which contained 339 patients, comparing pancreatic cancer vs. healthy controls [5].

Within these datasets, we built learning curves (Supplementary Section B), gathered relevant dataset-level characteristics, and measured the sample size where AUC stability was reached. We then compared the sample sizes measured directly from the learning curves to predicted sample sizes obtained using only our models through mean absolute error (MAE) and width of the 95% confidence intervals. We also recorded the AUC reached at the predicted sample sizes and compared this measure to the expected AUCs. As a sensitivity analysis, we swapped the three TCGA datasets for the two GEO datasets and re-fit models, again comparing validation performance and consistency of parameter estimates (Supplementary Section C).

### Comparison of Results to Traditional Power Analysis

Traditional sample size calculation methods for RNA-Seq data focus on calculating required *n* needed to confidently detect effects in a differential analysis, or univariable comparison of mean expression levels across genes and outcome classes. ML approaches, however, focus on the combined contribution of a gene set to classify the outcome effectively. Thus, we aimed to explore and compare the results obtained from this study to established power analysis methods for RNA-Seq data. Specifically, we compared required sample sizes for ML among the two validation datasets to sample sizes calculated using the *ssizeRNA* package [16]. There are many different tools available for calculating differential expression power and sample sizes; however, we opted to choose *ssizeRNA* as it has recently been shown to be the most accurate [12]. Further details can be found in Supplementary Section D. We discuss differences and similarities in sample sizes calculated between the two approaches and provide guidance on how both methods can be incorporated into future study design.

## Data Availability

All datasets used in this study are publicly available and can be found at https://www.ncbi.nlm.nih.gov/geo/. Furthermore, R Code used for further pre-processing and analysis is available from the corresponding author on request. Following acceptance, an R Shiny app will be made publicly available, where users can input dataset-level variables and receive training set size recommendations based on the results of this study.

## Data Availability

All datasets used in this study are publicly available and can be found at
https://www.ncbi.nlm.nih.gov/geo/. Furthermore, R Code used for further pre-processing and
analysis is available from the corresponding author on request. Following acceptance, an R
Shiny app will be made publicly available, where users can input dataset-level variables and
receive training set size recommendations based on the results of this study.

## References

1. Perakakis N, Yazdani A, Karniadakis GE, Mantzoros C. Omics, big data and machine learning as tools to propel understanding of biological mechanisms and to discover novel diagnostics and therapeutics. Metabolism. 2018 Oct;87:A1–9.

2. Holt RA, Jones SJM. The new paradigm of flow cell sequencing. Genome Research. 2008 May 7;18(6):839–46.

3. Smail C, Montgomery SB. RNA Sequencing in Disease Diagnosis. Annual Review of Genomics and Human Genetics [Internet]. 2024 Feb 15 [cited 2024 Oct 10];25(1). Available from: https://pubmed.ncbi.nlm.nih.gov/38360541/

4. Jiang F, Jiang Y, Zhi H, Dong Y, Li H, Ma S, et al. Artificial intelligence in healthcare: past, present and future. Stroke Vasc Neurol. 2017;2(4):230–243.

5. Yu S, Li Y, Liao Z, Wang Z, Wang Z, Li Y, et al. Plasma extracellular vesicle long RNA profiling identifies a diagnostic signature for the detection of pancreatic ductal adenocarcinoma. Gut. 2019 Sep 27;69(3):540–50.

6. Koch CM, Chiu SF, Akbarpour M, Bharat A, Ridge KM, Bartom ET, et al. A Beginner’s Guide to Analysis of RNA Sequencing Data. American Journal of Respiratory Cell and Molecular Biology. 2018 Aug 1;59(2):145–57.

7. Church BV, Williams HT, Mar JC. Investigating skewness to understand gene expression heterogeneity in large patient cohorts. BMC Bioinformatics. 2019 Dec;20(S24).

8. Teufel M, Sobetzko P. Reducing costs for DNA and RNA sequencing by sample pooling using a metagenomic approach. BMC Genomics [Internet]. 2022 Aug 24 [cited 2022 Nov 13];(1):1–10.

9. Jones C, Gannon B, Wakai A, O’Sullivan R. A systematic review of the cost of data collection for performance monitoring in hospitals. Systematic Reviews. 2015 Apr 1;4(1).

10. Mallik S, Zhao Z. Identification of gene signatures from RNA-seq data using Pareto-optimal cluster algorithm. BMC Systems Biology. 2018 Dec;12(S8).

11. Cheng Y, Xu SM, Santucci K, Lindner G, Janitz M. Machine learning and related approaches in transcriptomics. Biochemical and Biophysical Research Communications. 2024 Sep 1;724:150225–5.

12. Jeon H, Xie J, Jeon Y, Kyeong Joo Jung, Gupta A, Chang W, et al. Statistical Power Analysis for Designing Bulk, Single-Cell, and Spatial Transcriptomics Experiments: Review, Tutorial, and Perspectives. Biomolecules [Internet]. 2023 Jan 24 [cited 2023 May 1];13(2):221–1. Available from: https://www.ncbi.nlm.nih.gov/pmc/articles/PMC9952882/

13. Tomasz Burzykowski, Geubbelmans M, Rousseau AJ, Valkenborg D. Validation of machine learning algorithms. American Journal of Orthodontics and Dentofacial Orthopedics. 2023 Aug 1;164(2):295–7.

14. Gallitto G, Englert R, Kincses B, Kotikalapudi R, Li J, Hoffschlag K, et al. External validation of machine learning models—registered models and adaptive sample splitting. GigaScience [Internet]. 2025 [cited 2025 Aug 14];14. Available from: https://academic.oup.com/gigascience/article/doi/10.1093/gigascience/giaf036/8131472

15. Argmann C, Hou R, Ungaro RC, Irizar H, Al-Taie Z, Huang R, et al. Biopsy and blood-based molecular biomarker of inflammation in IBD. Gut. 2022 Sep 15;gutjnl-2021-326451.

16. Bi R, Liu P. Sample size calculation while controlling false discovery rate for differential expression analysis with RNA-sequencing experiments. BMC Bioinformatics. 2016 Mar 31;17(1).

17. Antunes-Ferreira M, D’Ambrosi S, Arkani M, Post E, In ‘t Veld SGJG, Ramaker J, et al. Tumor-educated platelet blood tests for Non-Small Cell Lung Cancer detection and management. Scientific Reports [Internet]. 2023 Jun 8;13(1):9359. Available from: https://www.nature.com/articles/s41598-023-35818-w

18. Hart SN, Therneau TM, Zhang Y, Poland GA, Pierre J. Calculating Sample Size Estimates for RNA Sequencing Data. Journal of Computational Biology. 2013 Nov 21;20(12):970–8.

19. Burcu Vitrinel, Koh L, Funda Mujgan Kar, Shuvadeep Maity, Rendleman J, Choi H, et al. Exploiting Interdata Relationships in Next-generation Proteomics Analysis. Molecular & Cellular Proteomics. 2019 Aug 9;18(8):S5–14.

20. Home - GEO - NCBI [Internet]. Nih.gov. 2019. Available from: https://www.ncbi.nlm.nih.gov/geo/

21. National Cancer Institute. The Cancer Genome Atlas Program (TCGA) - NCI [Internet]. www.cancer.gov. 2022. Available from: https://www.cancer.gov/ccg/research/genome-<u>sequencing/tcga</u>

22. Sanchis P, Lavignolle R, Abbate M, Lage-Vickers S, Vazquez E, Cotignola J, et al. Analysis workflow of publicly available RNA-sequencing datasets. STAR Protocols. 2021 Jun;2(2):100478.

23. Thennavan A, Beca F, Xia Y, Garcia-Recio S, Allison K, Collins LC, et al. Molecular analysis of TCGA breast cancer histologic types. Cell Genomics [Internet]. 2021 Dec 8 [cited 2022 Sep 1];1(3):100067.

24. Anusewicz D, Orzechowska M, Bednarek AK. Lung squamous cell carcinoma and lung adenocarcinoma differential gene expression regulation through pathways of Notch, Hedgehog, Wnt, and ErbB signalling. Scientific Reports. 2020 Dec;10(1).

25. Захарова ГС, Efimov V, Mikhail Raevskiy, Румянцев ПО, Gudkov A, Oksana Yu. Belogurova-Ovchinnikova, et al. Reclassification of TCGA Diffuse Glioma Profiles Linked to Transcriptomic, Epigenetic, Genomic and Clinical Data, According to the 2021 WHO CNS Tumor Classification. International Journal of Molecular Sciences. 2022 Dec 21;24(1):157–7.

26. Brennan Cameron W, Verhaak Roel GW, McKenna A, Campos B, Noushmehr H, Salama Sofie R, et al. The Somatic Genomic Landscape of Glioblastoma. Cell. 2013 Oct;155(2):462–77.

27. Akshata Desai KA. Triple Negative Breast Cancer – An Overview. Hereditary Genetics [Internet]. 2012; Available from: https://www.ncbi.nlm.nih.gov/pmc/articles/PMC4181680/

28. Devijver P, Kittler J. Pattern Recognition: A Statistical Approach. New Jersey, United States. Prentice-Hall; 1982.

29. Webb GI, Sammut C, Perlich C, Horváth T, Wrobel S, Korb KB. Learning curves in machine learning. Encyclopedia of Machine Learning. 2011:577–580.

30. Hanley JA, McNeil BJ. The meaning and use of the area under a receiver operating characteristic (ROC) curve. Radiology. 1982;143(1):29–36.

31. Bradley AP. The use of the area under the ROC curve in the evaluation of machine learning algorithms. Pattern Recognition. 1997;30(7):1145–1159.

32. Kaur D, Sobiesk M, Patil S, Liu J, Bhagat P, Gupta A, et al. Application of Bayesian networks to generate synthetic health data. Journal of the American Medical Informatics Association: JAMIA [Internet]. 2021 Mar 18 [cited 2022 Nov 16];28(4):801–11. Available from: https://pubmed.ncbi.nlm.nih.gov/33367620/

33. Love MI, Huber W, Anders S. Moderated estimation of fold change and dispersion for RNA-seq data with DESeq2. Genome Biology. 2014 Dec;15(12):550.

34. Kursa MB, Rudnicki WR. Feature Selection with theBorutaPackage. Journal of Statistical Software. 2010;36(11).

35. Wright MN, Ziegler A. Ranger: a fast implementation of random forests for high dimensional data in C++ and R. J. Stat. Soft. 2017;77(1):1–17.

36. Chen T, Guestrin C. XGBoost: a scalable tree boosting system. 2016. Presented at: Proceedings of the 22nd ACM SIGKDD International Conference on Knowledge Discovery and Data Mining - KDD-16; August 13-17, 2016:785-794; San Francisco, California, USA.

37. Mall PK, Singh PK, Srivastav S, Narayan V, Paprzycki M, Jaworska T, et al. A comprehensive review of deep neural networks for medical image processing: recent developments and future opportunities. Healthcare Analytics. 2023;4:100216.

38. Nwanosike EM, Conway BR, Merchant HA, Hasan SS. Potential applications and performance of machine learning techniques and algorithms in clinical practice: a systematic review. Int J Med Inform. 2022;159:104679.

39. Foundation for Open Access Statistics. Fast scalable R with H20. 2015. URL: https://h2o.ai/ [accessed 2024-11-25]

40. Silvey S, Liu J. Sample Size Requirements for Popular Classification Algorithms in Tabular Clinical Data: Empirical Study. Journal of Medical Internet Research. 2024 Dec 17;26:e60231.

41. Hilbe J. Negative Binomial Regression. Cambridge, England. Cambridge University Press; 2007.

42. Bozdogan H. Model selection and Akaike’s Information Criterion (AIC): the general theory and its analytical extensions. Psychometrika. 1987;52(3):345–370.

43. Vanegas L, Rondón L, Paula G. _glmtoolbox: Set of tools to data analysis using generalized linear models_. 2024. URL: https://CRAN.R-project.org/package=glmtoolbox [accessed 2024- 11-19]

44. Veall MR, Zimmermann KF. Pseudo-R2 measures for some common limited dependent variable models. Journal of Economic Surveys. 2006;10(3):241–259.

45. Wang Q, Armenia J, Zhang C, Penson AV, Reznik E, Zhang L, et al. Unifying cancer and normal RNA sequencing data from different sources. Scientific Data [Internet]. 2018 Apr 17 [cited 2019 Nov 27];5(1). Available from: https://www.nature.com/articles/sdata201861

