## Supplementary Material for "Sample Size Requirements for Machine Learning Classification of Binary Outcomes in Bulk RNA-Seq Data"

**Supplementary Section A: Bayesian Network Generation and Data Simulation Technique**

Bayesian Network Generation [1] is a statistical learning technique that aims to learn the structure of a set of variables and their conditional dependencies using Directed Acyclic Graphs (DAG). In the context of simulating RNA-Seq data, we can describe the process mathematically using the following components and formulas:

**1. Directed Acyclic Graph (DAG) Representation**

Let G = (V, E) be a directed acyclic graph where:

- V is a set of nodes representing the variables (e.g., genes or gene expression levels).
- E is a set of directed edges representing the dependencies between the variables.

**2. Joint Probability Distribution**

The joint probability distribution *P*(X_1_, X_2_…. X*_n_*) of the variables can be expressed using the chain rule of probability for Bayesian networks: *P*(X_1_, X_2_…. X*_n_*) = $\prod_{i=1}^{n} P(Xi| Pa(Xi))$ where $Pa(Xi)$ denotes the set of parents of node $Xi$ in the DAG.

**3.** **Conditional Probability Tables (CPTs)**

Each node $Xi$ has an associated conditional probability table (CPT) that defines the probability of $Xi$ given its parents: $P(Xi| Pa(Xi)) for all i.$

BNG approaches have been shown to produce data that is structurally similar to the original source [2]. The first step in building a Bayesian network is construction of a “skeleton,” or structure of the DAG, which specifies all meaningful conditional dependencies between genes. For example, let a simple network consist of three genes (A,B,C), with conditional probabilities of interest being *P*(B|A) and *P*(C|A), implying that the expression level of gene A influences genes B and C . This network can be visualized below:

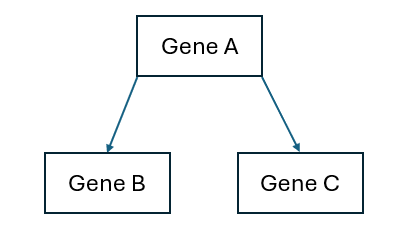

Once this structure is specified, simulating data from this (Gaussian) Bayesian network would begin by sampling from node A where *A* ∼ *N*(*μ*_A_,*σ*_A_^2^), which does not have any parents, and then sample from the conditional distributions *P*(B|A) and *P*(C|A), which will also be Normally distributed.

In a setting where there are many genes, selecting the most accurate skeleton is computationally expensive using an exhaustive approach; therefore, a variety of score-based and other combinatoric algorithms can be used to select an optimal graph. We used the “Tabu” algorithm to learn the structure of our non-simulated RNA-Seq datasets [3]. The Tabu algorithm uses local neighborhood searching to find the Bayesian Network with the best score function. Tabu was selected for use in this project due to its speed and reliability in selecting a Bayesian Network that adequately represents the original data source [4].

Our specific BNG simulation approach, tailored for RNA-Seq type data, was as follows:

(At this point, we assume that the differentially expressed gene set has already been determined, and the input here is DESeq2-normalized count values.)

1.) Separate outcome classes into two separate sets. Transform data in each outcome class to approximately Normal using log2(*X*+0.01) mapping.
2.) Select network skeleton using Tabu algorithm in each outcome class separately.
3.) Train Bayesian network on differentially expressed gene set within each outcome class separately using the “bn.fit” function in R [5].
4.) Simulate 5,000 total observations from the fitted network for each outcome class using “rbn” function in R [5]. For each outcome class, the number of observations simulated were equal to 5,000 times the original outcome proportion.
5.) Pool the simulated networks for each outcome class together.
6.) Final data is normally distributed [1]. Back-transform the final data into skewed pseudo-counts using 2*^X^*^-0.01^ mapping.

**Supplementary Section B:**

**Subsection 1:**

*Specific Learning Curve Strategy*

The specific learning curve strategy was as follows:

**In the simulated datasets (“GBM” to “MS”):**

1.) Create a list of proposed sample sizes *n* (See supp. table 1).
2.) At each point in the sample size interval, randomly sample 50 sub-datasets of size 2*n* from the full dataset. Half of the subsampled data is used for training, and the other half is reserved for testing (to ensure independence).
3.) In each training dataset, fit the machine learning model of choice and estimate AUC in the testing set. Average the 50 mean-validated AUC values to generate an estimate of out-of-sample performance at a given *n*.
4.) Repeat at the next *n* in the list.

**In the non-simulated datasets (TCGA-Brain, TCGA-Lung, TCGA-Breast, IBD, and PDAC):**

Prior to this, entire datasets were randomly separated into training and testing sets. The full dataset sizes were as follows: TCGA-Brain: 875. TCGA-Breast: 925. TCGA-Lung: 1162. IBD: 2471. PDAC: 339. Training set sizes were as follows: TCGA-Brain: 800. TCGA-Breast: 875. TCGA-Lung: 1000. IBD: 2000. PDAC: 300. Test set sizes were: TCGA-Brain: 75. TCGA-Breast: 50. TCGA-Lung: 162. IBD: 471. PDAC: 39.

1.) Create a list of proposed sample sizes *n* (See Subsection 2).
2.) At each point in the sample size interval, randomly sample 50 sub-datasets of size *n* from the full training dataset.
3.) In each sub-dataset, fit the machine learning model of choice and estimate AUC in the pre-selected testing set. Average the 50 mean-validated AUC values to generate an estimate of out-of-sample performance at a given *n*.
4.) Repeat at the next *n* in the list.

Once raw data was collected, estimated learning curves were fit using nonlinear least squares optimization [6], following the power law equation: *AUC_(n)_* = *an^b^* + *c*, where *a* and *b* were estimated, and *c* was either fixed to be the full-dataset AUC or was also estimated, depending on the visual quality of the fit. For some datasets and algorithms, the power law function did not fit the data well. In these scenarios, we instead fit the learning curves using a logarithmic function, *AUC_(n)_* = *β_0_* + *β_1_***log*(*n)*, where *β_0_* and *β_1_* were estimated using ordinary least-squares [7].

**Subsection 2:**

**Learning Curve Sample Size Lists and Fitting Methods Used**

*In order : number of points examined, range of potential sample sizes.

**In Order : XGB, RF, NN. 1 = Power Law (*c* estimated), 2 = Power Law (*c* fixed to full-dataset AUC), 3 = log-linear regression.

| **Dataset** | **XGB*** | **RF*** | **NN*** | **Curve Fitting Method**** |
| --- | --- | --- | --- | --- |
| GBM | 10, [50-250] | 10, [50-250] | 10, [50-250] | 1,1,2 |
| HCC | 10, [50-250] | 10, [50-250] | 10, [50-500] | 1,1,1 |
| Kidney | 10, [50-1000] | 10, [50-1000] | 10, [50-2000] | 3,3,2 |
| IPF | 10, [25-150] | 10, [25-150] | 10, [25-250] | 2,3,2 |
| COVID | 10, [50-2000] | 10, [50-2000] | 10, [50-2000] | 1,1,3 |
| HBV | 10, [50-1000] | 10, [50-1000] | 10, [100-2000] | 3,3,2 |
| Hypertension | 10, [50-250] | 10, [50-250] | 10, [50-500] | 2,2,2 |
| NAFLD | 10, [50-250] | 10, [50-250] | 10, [50-500] | 1,2,1 |
| PrePost | 10, [50-500] | 10, [50-500] | 10, [50-500] | 1,1,1 |
| RA | 10, [50-750] | 10, [50-750] | 10, [50-2000] | 1,1,3 |
| AVSC | 10, [50-1500] | 10, [50-1500] | 10, [50-2000] | 1,1,1 |
| Crohn | 10, [50-2000] | 10, [50-2000] | 10, [50-2000] | 1,1,3 |
| MDD | 10, [50-2000] | 10, [50-2000] | 10, [50-2000] | 3,1,2 |
| Ovarian | 10, [50-2000] | 10, [50-2000] | 10, [50-2000] | 1,1,3 |
| ALS | 10, [50-2000] | 10, [50-2000] | 10, [50-2000] | 1,1,1 |
| Kawasaki | 10, [50-250] | 10, [50-250] | 10, [50-2000] | 2,2,1 |
| CCA | 10, [50-1000] | 10, [50-1000] | 10, [50-2000] | 1,1,1 |
| Glioma | 10, [50-1000] | 10, [50-1000] | 10, [50-2000] | 1,1,3 |
| TBProg | 10, [50-2000] | 10, [50-2000] | 10, [50-2000] | 1,1,1 |
| Tuberculosis | 10, [75-500] | 10, [75-500] | 10, [50-500] | 2,1,2 |
| EDS | 10, [50-250] | 10, [50-250] | 10, [50-250] | 1,1,1 |
| NSCLC | 10, [50-500] | 10, [50-500] | 10, [50-1000] | 1,1,1 |
| Bipolar | 10, [50-2000] | 10, [50-2000] | 10, [50-2000] | 1,1,1 |
| MS | 10, [50-2000] | 10, [50-2000] | 10, [50-2000] | 1,1,3 |
| TCGA-Brain | 10, [25-250] | 10, [25-250] | 10, [25-250] | 1,2,1 |
| TCGA-Breast | 10, [25-800] | 10, [25-800] | 10, [25-800] | 2,3,3 |
| TCGA-Lung | 10, [25-250] | 10, [25-250] | 10, [25-250] | 1,2,1 |
| **Validation Datasets** |  |  |  |  |
| PDAC | 10, [50-290] | 10, [50-290] | 10, [50-290] | 1,1,2 |
| IBD | 10, [50-1500] | 10, [50-1500] | 10, [50-1500] | 1,1,2 |

**Subsection 3:**

**Full Table of Dataset-Level Characteristics and Their Definitions:**

| **Dataset Characteristic** | **Definition** |
| --- | --- |
| Minority Class Proportion | Class imbalance. The percentage of observations corresponding to the minority outcome class. |
| Number of Differentially Expressed Genes | Number of genes that were both selected by DESeq2 (FDR-adjusted *p*-value < 0.05) and Boruta algorithm. |
| Median LFC | The median absolute log_2_-tranformed fold-change between the outcome groups, among the differentially expressed genes. |
| Maximum LFC | The maximum absolute log_2_-tranformed fold-change between the outcome groups, among the differentially expressed genes. |
| Minimum LFC | The minimum absolute log_2_-tranformed fold-change between the outcome groups, among the differentially expressed genes. |
| Dispersion | The median estimated dispersion parameter among the differentially expressed genes. Dispersion calculated using DESeq2 definition. In final models, categorized as “high” > 1, vs. “low” < 1. |
| Correlation | The median absolute inter-gene correlation coefficient among the differentially expressed genes. Calculated from the upper diagonal of the log_2_-transformed correlation matrix of the predictor set. In final models, categorized as “high” > 0.5, vs. “low” < 0.5. |
| Median Avg. Read Count | The median average read count among the differentially expressed genes. Averaged read counts calculated using DESeq2-normalized counts. |
| Separability | The maximum AUC attained by each machine-learning method, in the complete simulated dataset. Provides an estimate of potential asymptotic performance. In final models, categorized as “high” > 0.99, vs. “low” < 0.99. |
| Nonlinearity | Difference in maximum-achievable AUC scores between the best-performing machine learning algorithm, compared to a standard multivariable logistic regression model. In final models, categorized as “high” > 4.5-point difference, vs. “low” < 4.5-point difference (see Supplementary Section F). |

**Subsection 4:**

**Sample Sizes Needed to Detect AUC within 2-points (0.02) of Maximum Achievable Value (Main Analysis):**

| *Dataset Name* | *XGB* | *RF* | *NN* |
| --- | --- | --- | --- |
| GBM | 54 | 50 | 50 |
| HCC | 168 | 128 | 324 |
| Kidney | 813 | 152 | 227 |
| IPF | 81 | 25 | 25 |
| COVID | 631 | 400 | 1,374 |
| HBV | 529 | 106 | 144 |
| Hypertension | 126 | 56 | 212 |
| NAFLD | 72 | 50 | 50 |
| PrePost | 154 | 93 | 199 |
| RA | 455 | 344 | 761 |
| AVSC | 1,083 | 533 | 888 |
| Crohn | 613 | 191 | 681 |
| MDD | 950 | 263 | 156 |
| Ovarian | 713 | 215 | 565 |
| ALS | 1,048 | 970 | 1,537 |
| Kawasaki | 60 | 50 | 57 |
| CCA | 642 | 473 | 960 |
| Glioma | 480 | 353 | 1,047 |
| TBProg | 904 | 1,005 | 1,444 |
| Tuberculosis | 270 | 357 | 239 |
| EDS | 132 | 50 | 94 |
| NSCLC | 235 | 189 | 371 |
| Bipolar | 886 | 644 | 1,471 |
| MS | 1,200 | 1,307 | 1,711 |
| TCGA-Brain | 68 | 25 | 25 |
| TCGA-Breast | 607 | 419 | 298 |
| TCGA-Lung | 86 | 25 | 69 |
| *Median (Range)* | 468 (54-1,200) | 190 (25-1,307) | 269 (25-1,711) |

**Sample Sizes at Alternative Thresholds of 0.01 and 0.05:**

| **Threshold: 0.01** | | | |
| --- | --- | --- | --- |
| *Dataset Name* | *XGB* | *RF* | *NN* |
| GBM | 87 | 42 | 51 |
| HCC | 303 | 531 | >2,500 |
| Kidney | 1,619 | 918 | 706 |
| IPF | 175 | <25 | <25 |
| COVID | 1,376 | 881 | 1,792 |
| HBV | 1,138 | 463 | 404 |
| Hypertension | 279 | 142 | 777 |
| NAFLD | 134 | 52 | 84 |
| PrePost | 360 | 222 | >2,500 |
| RA | 907 | 795 | 1,542 |
| AVSC | >2,500 | 1,275 | 1,361 |
| Crohn | 1,311 | 404 | 1,325 |
| MDD | 1,536 | 654 | 354 |
| Ovarian | 1,377 | 639 | 1,810 |
| ALS | 1,763 | 1,752 | 2,265 |
| Kawasaki | 120 | 33 | 92 |
| CCA | 1,034 | 943 | 2,012 |
| Glioma | 841 | 772 | 2,148 |
| TBProg | 1,591 | 1,797 | >2,500 |
| Tuberculosis | 483 | 831 | 695 |
| EDS | 209 | 109 | 180 |
| NSCLC | 450 | 463 | 806 |
| Bipolar | 1,707 | 1,518 | >2,500 |
| MS | 1,780 | >2,500 | 2,068 |
| TCGA-Brain | 112 | <25 | <25 |
| TCGA-Breast | >2,500 | 686 | 485 |
| TCGA-Lung | 195 | 83 | 167 |
| *Median (Range)* | 907 [87-2,500] | 639 [25-2,500] | 806 [25-2,500] |
| **Threshold: 0.05** | | | |
| *Dataset Name* | *XGB* | *RF* | *NN* |
| GBM | 30 | <25 | 29 |
| HCC | 45 | <25 | 54 |
| Kidney | 103 | <25 | 51 |
| IPF | 29 | <25 | <25 |
| COVID | 157 | 94 | 619 |
| HBV | 53 | <25 | 37 |
| Hypertension | 44 | <25 | 38 |
| NAFLD | 28 | <25 | <25 |
| PrePost | 53 | 35 | 48 |
| RA | 122 | 64 | 92 |
| AVSC | 196 | 136 | 383 |
| Crohn | 148 | 71 | 93 |
| MDD | 225 | 40 | 53 |
| Ovarian | 164 | <25 | <25 |
| ALS | 360 | 280 | 621 |
| Kawasaki | <25 | <25 | 38 |
| CCA | 202 | 102 | 178 |
| Glioma | 137 | 53 | 122 |
| TBProg | 286 | 301 | 482 |
| Tuberculosis | 125 | 96 | 58 |
| EDS | 58 | <25 | 44 |
| NSCLC | 71 | 32 | 96 |
| Bipolar | 247 | 125 | 369 |
| MS | 477 | 365 | 968 |
| TCGA-Brain | 30 | <25 | <25 |
| TCGA-Breast | 69 | 96 | 69 |
| TCGA-Lung | 40 | <25 | <25 |
| *Median (Range)* | 103 [25-477] | 35 [25-365] | 54 [25-986] |

**Supplementary Section C:** Model Validation on Swapped Data

As mentioned in the main text, our presented sample size models used a combination of simulated and non-simulated experimental data (24 GEO + 3 TCGA, reserving two GEO datasets for validation). In order to ensure that this was not biased, we re-fit models by including IBD and PDAC and omitting TCGA-Lung, Brain, and Breast.

Re-fit model estimates are as follows:

Validation results of the re-fit model on the TCGA datasets can be seen below.

*XGBoost Model*:

Exponentiated Coefficient Estimates:

Dispersion: 1.33 (vs. 1.38 in original model)

Separability: 0.23 (vs. 0.25)

Min. LFC: 1.49 (vs. 1.66)

Med. LFC: 0.61 (vs. 0.56)

Adj. R^2^: 0.82 (vs. 0.84)

*Random Forest Model*:

Exponentiated Coefficient Estimates:

Max. LFC: 0.71 (vs. 0.72 in original model)

Dispersion: 1.64 (vs. 1.61)

Imbalance: 0.98 (vs. 0.99)

Nonlinearity: 3.43 (vs. 3.31)

Adj. R^2^: 0.83 (vs. 0.79)

*Neural Network Model*:

Exponentiated Coefficient Estimates:

Nonlinearity: 2.25 (vs. 2.44 in original model)

Separability: 0.45 (vs. 0.50)

Max. LFC: 0.81 (vs. 0.75)

Dispersion: 2.20 (vs. 2.33)

Adj. R^2^: 0.67 (vs. 0.69)

**Supplementary Section D:** Further Validation Data / Calculation of Differential Expression Sample Sizes

**Subsection 1:**

**Other Validation Dataset Characteristics:**

| *Dataset Name* | *Minority Class Proportion* | *Number of Differentially Expressed Genes* | *Median LFC (Min, Max)* | *Median Between-Gene Correlation* | *Median Dispersion* | *Median Avg. Read Count* |
| --- | --- | --- | --- | --- | --- | --- |
| PDAC | 30.1% | 67 | 0.87 (0.01, 5.58) | 0.32 | 0.89 | 5465.65 |
| IBD | 18.1% | 57 | 0.90 (0.17, 2.62) | 0.25 | 1.07 | 180.74 |

Sample sizes for traditional differential expression analysis were calculated using the *ssizeRNA* package [8]. As discussed in the main text, a recent review article published by Jeon et. al (2023) [9] cite two studies [8,10] that used simulation approaches to show that this method was the most accurate in obtaining the correct sample size. In order to calculate *n* from this package, the total number of potential genes, proportion of those expected to be differentially expressed, number of iterations for simulation approach, average read count/dispersion/fold-change for the genes of interest, FDR, and desired power.

Calculating simultaneous power for a gene panel with differing fold-changes, dispersions, etc. is complicated. Therefore, for each validation dataset (IBD and PDAC), we opted to use values of read count/dispersion/fold-change for the gene at the median fold-change value for all potential differentially expressed genes for simplicity, an FDR of 0.05, and desired power of 80%. Specifically, for IBD, the gene with median fold-change had an effect size of 1.87, read count of 39.48, and dispersion of 0.958. We also used: total genes 39,354, differentially expressed gene percentage: 57/39,354. For PDAC, the gene with median fold-change had an effect size of 1.83, read count of 13069.69, and dispersion of 0.209. We also used: total genes 35,923, differentially expressed gene percentage: 67/35,923.

**Subsection 2:**

**Validation of Re-Fit Model on Held-Out TCGA Datasets:**

| **Dataset** | ***XGBoost*** | | | |
| --- | --- | --- | --- | --- |
|  | **True *N*** | **Predicted *N* [95% CI]** | **Difference (Predicted – True)** | **AUC at Predicted *n*** |
| TCGA-Brain | 68 | 63* [31-128] | -5 | 0.978 (Expected: 0.980) |
| TCGA-Breast | 607 | 648* [498-841] | +41 | 0.699 (Expected: 0.698) |
| TCGA-Lung | 86 | 149* [40-556] | +63 | 0.973 (Expected: 0.965) |
|  | ***Random Forest*** | | | |
|  | **True *N*** | **Predicted *N*** | **Difference (Predicted – True)** | **AUC at Predicted *n*** |
| TCGA-Brain | 25 | 28* [16-50] | +3 | 0.994 (Expected: 0.980) |
| TCGA-Breast | 419 | 145 [110-190] | -274 | 0.709 (Expected: 0.730) |
| TCGA-Lung | 25 | 12* [5-27] | -13 | 0.958 (Expected: 0.963) |
|  | ***Neural Network*** | | | |
|  | **True *N*** | **Predicted *N*** | **Difference (Predicted – True)** | **AUC at Predicted *n*** |
| TCGA-Brain | 25 | 68 [38-122] | +43 | 0.994 (Expected: 0.980) |
| TCGA-Breast | 298 | 413* [273-626] | +115 | 0.711 (Expected: 0.704) |
| TCGA-Lung | 69 | 65* [25-166] | -4 | 0.960 (Expected: 0.961) |

*****True *n* is included in 95% confidence interval.

**Supplementary Section E:** Simulation Consistency Results

We plotted the average log-fold changes calculated over all features in each simulated data set compared to the log-fold changes arising from the original source. As seen below, the simulation procedure was able to closely resemble the original experimental datasets.

Median LFC (simulated data) vs. Median LFC (original source)

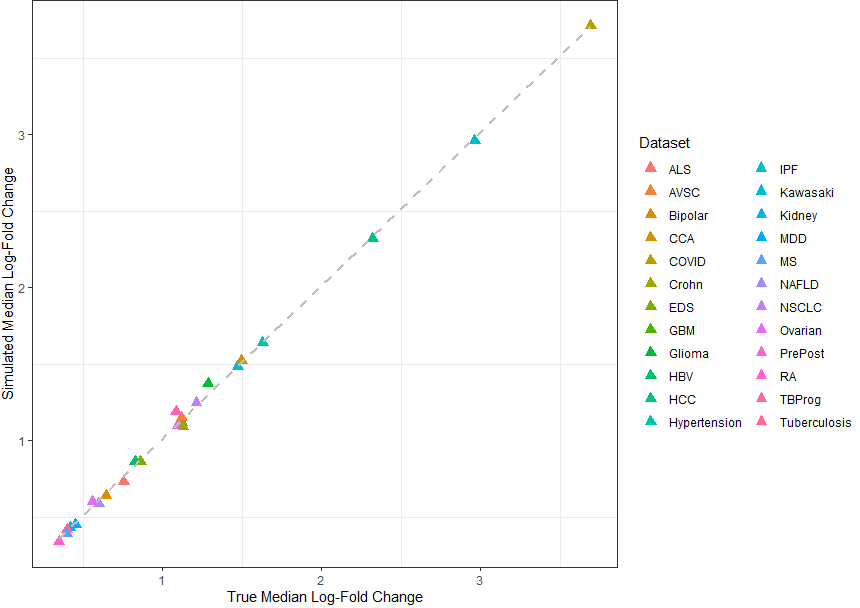

Min LFC (simulated data) vs. Min LFC (original source)

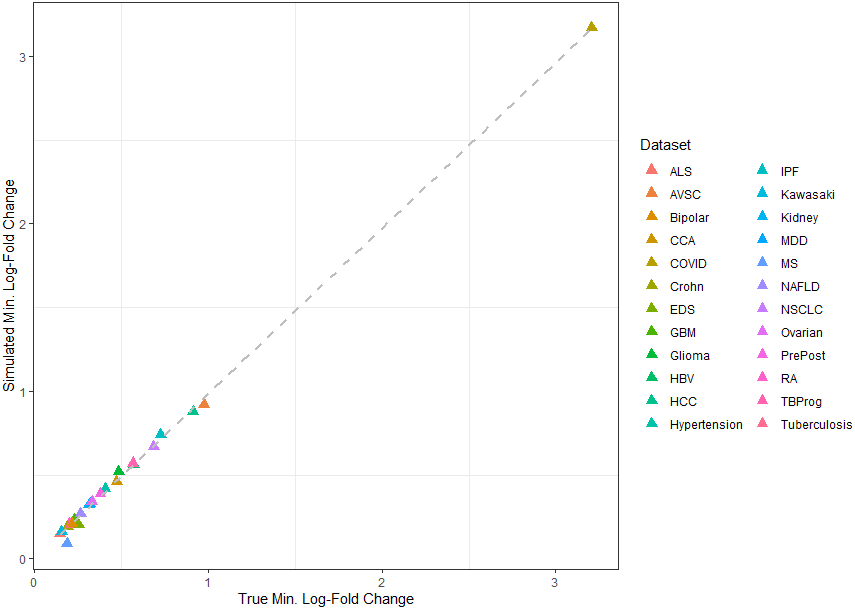

Max LFC (simulated data) vs. Max LFC (original source)

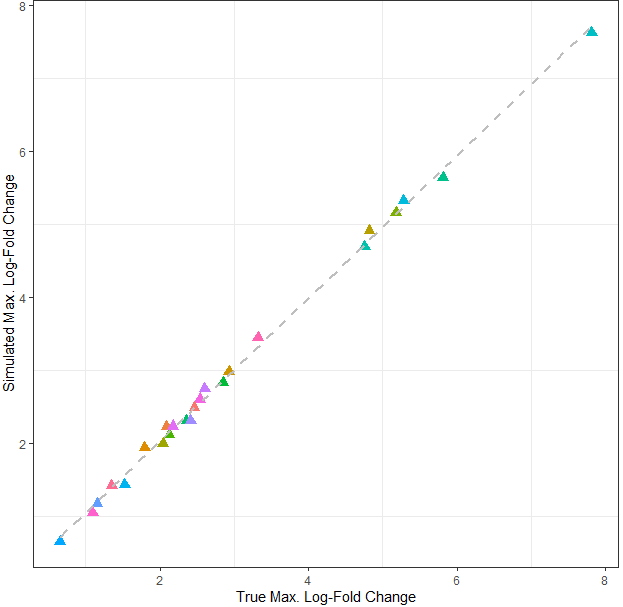

**Supplementary Section F:** Derivation of Cutoff Used to Categorize Dataset Nonlinearity Values

We examined a range of nonlinearity cutoffs ranging from 2 to 8.

Then, in univariable negative binomial regression models, we fit models using nonlinearity as a single predictor, and sample sizes as the response over XGB, RF, and NN algorithms separately. At each cutoff, we calculated the leave-one-out cross-validation mean squared error (LOO-CV MSE). A lower score indicates that the nonlinearity threshold was able to predict *n* with better precision. We then selected the lowest threshold for each algorithm based on results.

**Subsection 1:**

RF Results

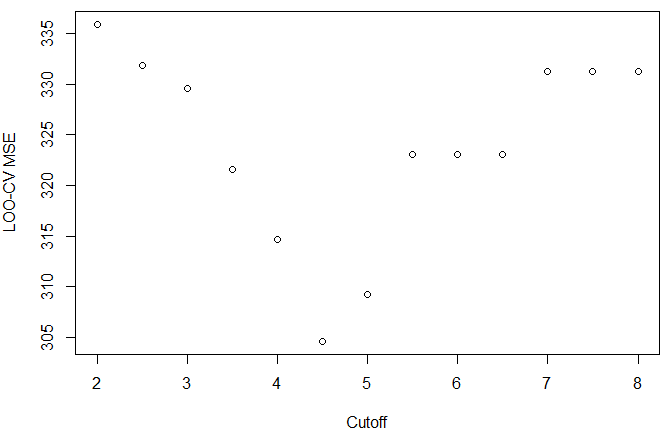

**Subsection 2:**

NN Results

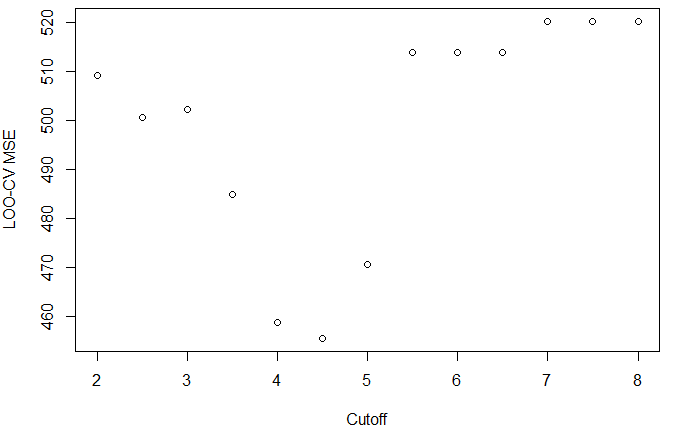

**Subsection 3:** XGB Results

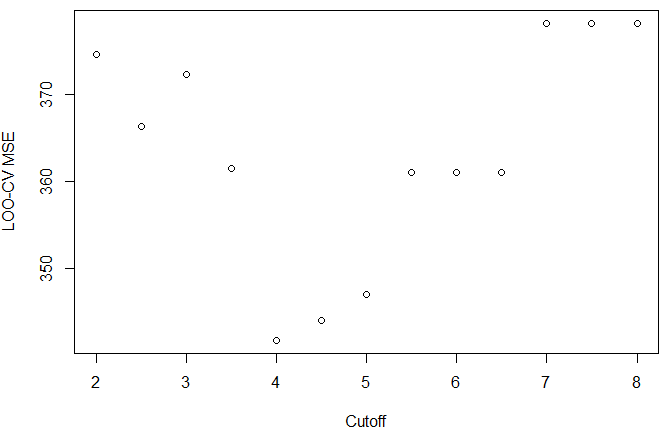

For RF and NN, a cutoff of 4.5 minimized LOO-CV MSE. For XGB, a cutoff of 4.0 minimized LOO-CV MSE. We elected to choose one unified cutoff of all three algorithms; therefore we chose 4.5 for downstream modelling.

**Supplementary Section G:**

**Subsection 1:** Visualization, Univariable Associations Between Dataset Characteristics and Sample Sizes (Max. AUC, nonlinearity, and max./median LFC are included in main text)

**Minimum LFC:**

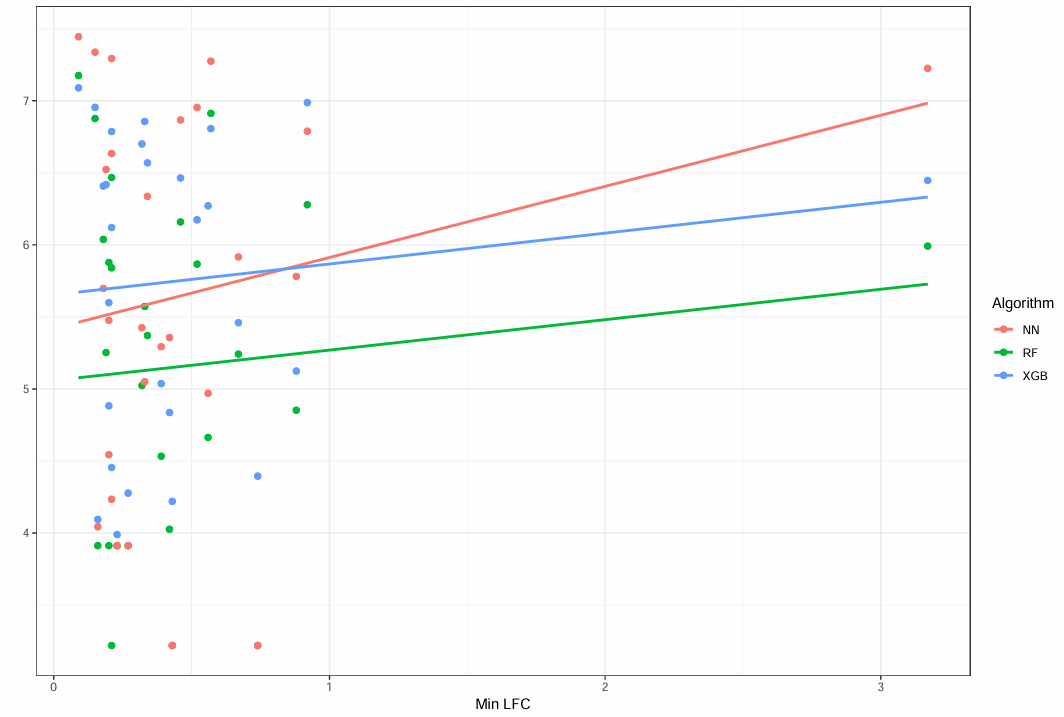

**Median Between-Gene Correlation:**

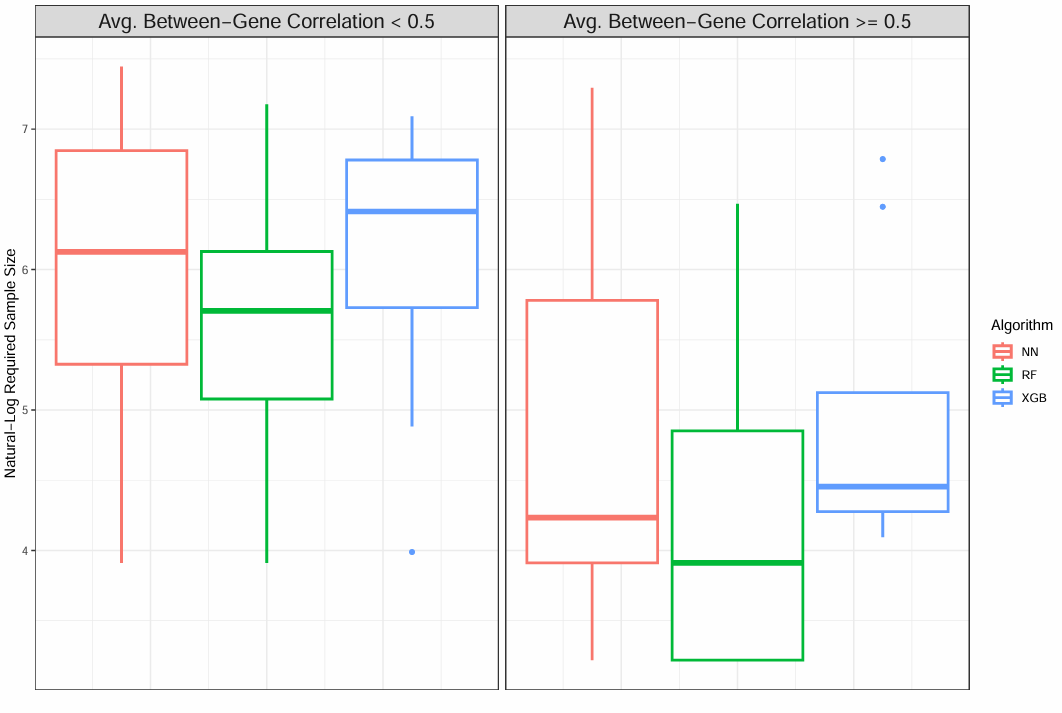

**Number of Genes included in ML modelling:**

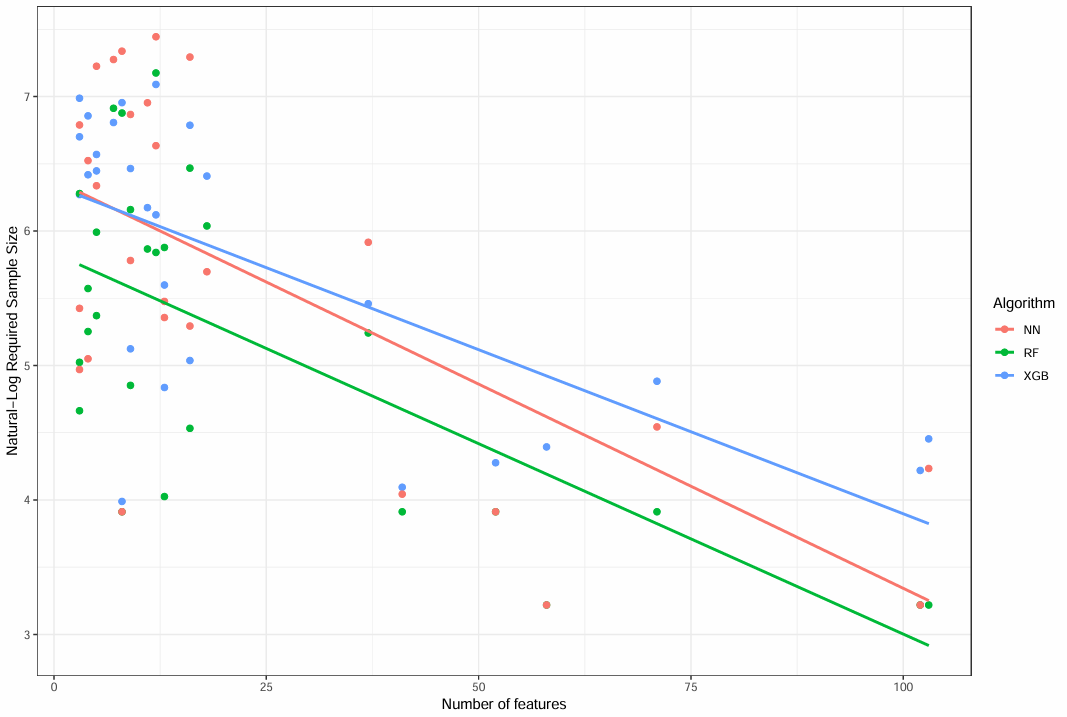

**Median Average Read Count:**

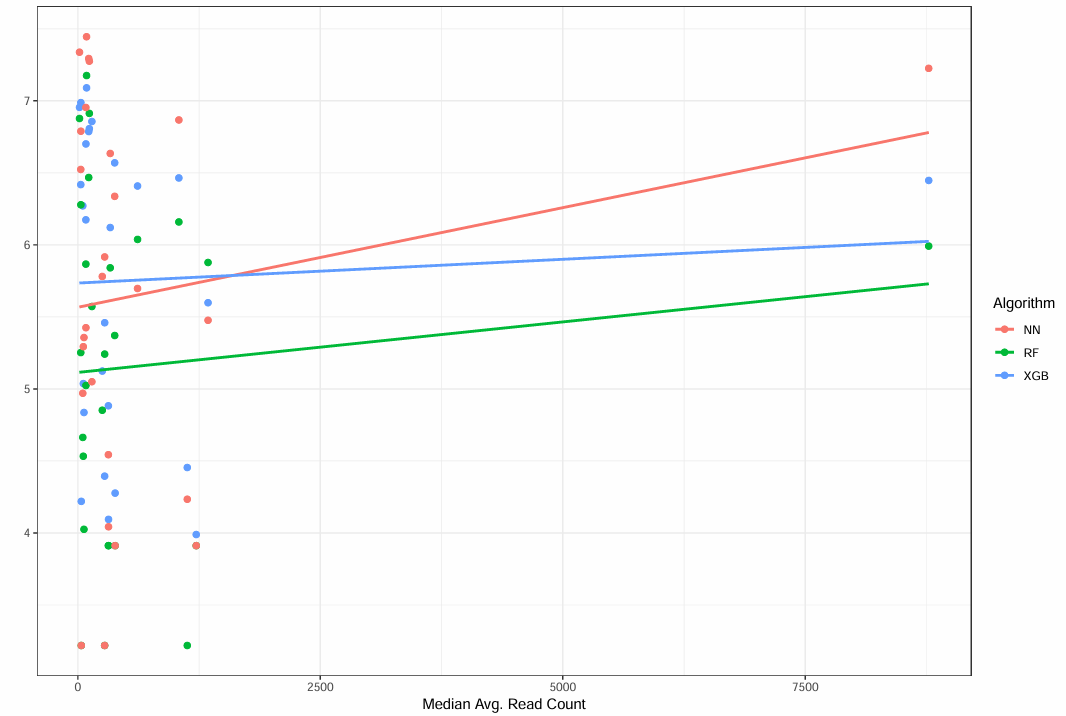

**Class Imbalance:**

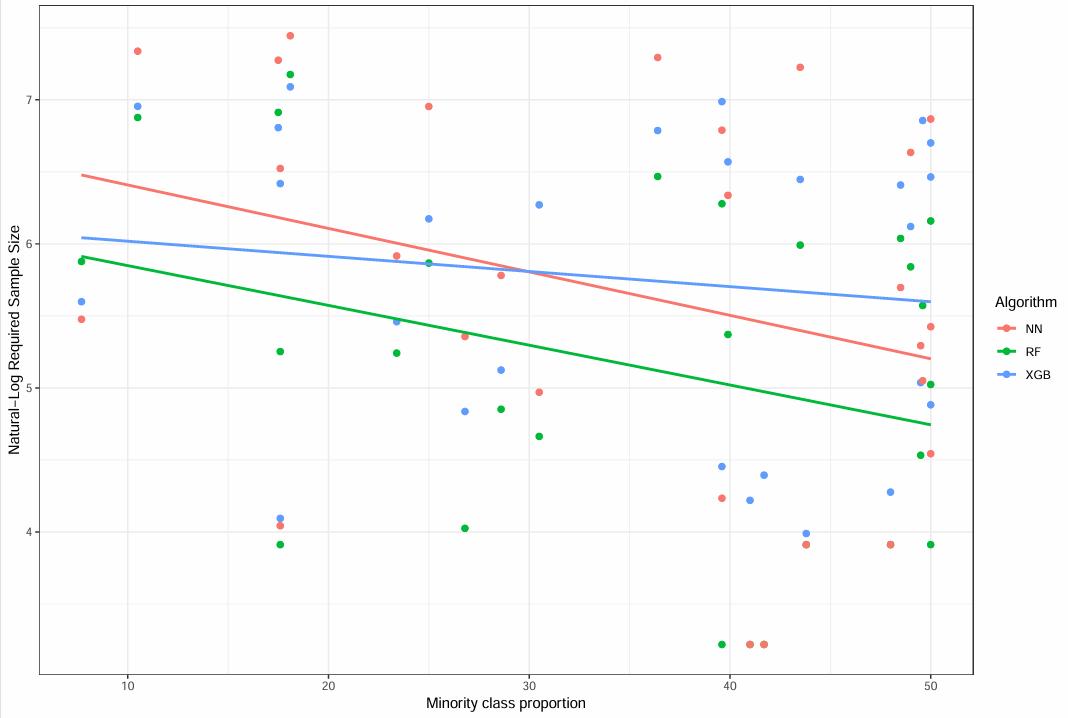

**Median Dispersion:**

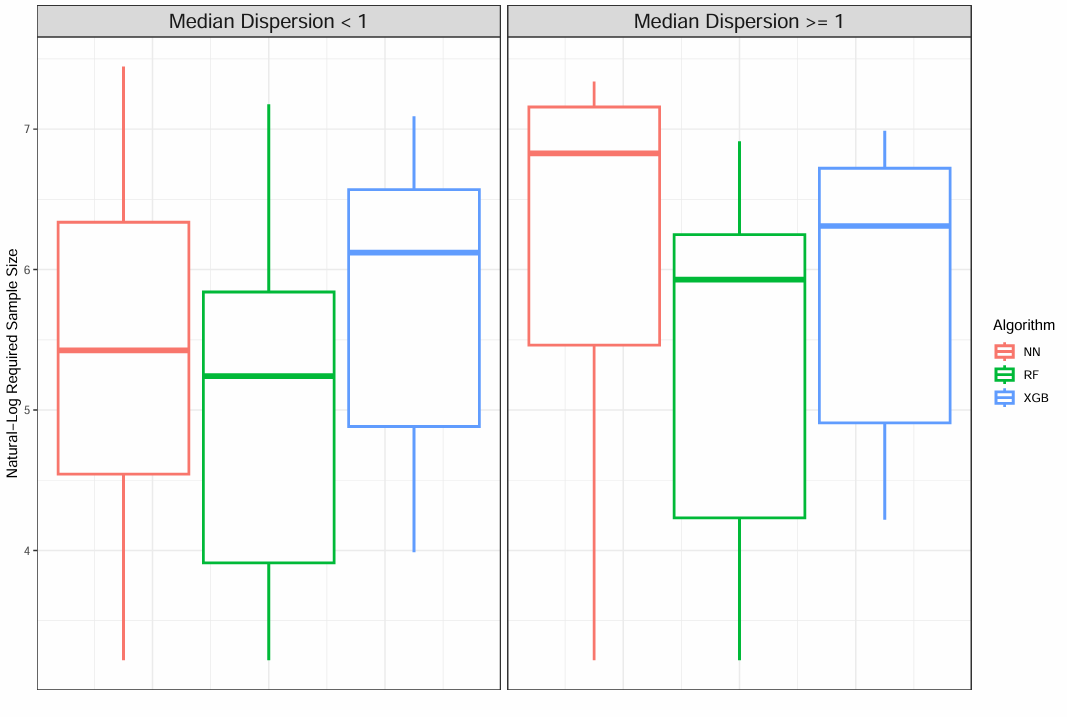

**Subsection 2:** Tables Detailing Univariable and Multivariable Negative Binomial Regression Models

**Univariable Associations between Dataset-Characteristics and Sample Sizes:**

| *Variable* | *XGB* | *RF* | *NN* |
| --- | --- | --- | --- |
| Minority Class Proportion | 0.992 [0.969, 1.015], *p*=0.537 | 0.973 [0.948, 0.997], ***p*=0.044** | 0.976 [0.947, 1.004], *p*=0.094 |
| Separability | 0.177 [0.122, 0.261], ***p*<0.001** | 0.211 [0.116, 0.393], ***p*<0.001** | 0.180 [0.097, 0.342], ***p*<0.001** |
| Number of Differentially Expressed Genes | 0.975 [0.968, 0.983], ***p*<0.001** | 0.968 [0.959, 0.979], ***p*<0.001** | 0.968 [0.959, 0.980], ***p*<0.001** |
| Average Read Count | 1.000 [0.999-1.000], *p*=0.986 | 1.000 [0.999-1.000], *p*=0.977 | 1.000 [0.999-1.000], *p*=0.542 |
| Dispersion | 1.138 [0.585, 2.303], *p*=0.709 | 1.497 [0.701, 3.373], *p*=0.308 | 1.887 [0.861, 4.376], *p*=0.121 |
| Correlation | 0.400 [0.216, 0.774], ***p*=0.005** | 0.397 [0.191, 0.879], ***p*=0.016** | 0.634 [0.282, 1.545], *p*=0.287 |
| Median LFC | 0.698 [0.542, 0.935], ***p*=0.014** | 0.666 [0.493, 0.951], ***p*=0.018** | 0.849 [0.604, 1.279], *p*=0.391 |
| Minimum LFC | 1.059 [0.672, 2.063], *p*=0.846 | 0.985 [0.608, 2.041], *p*=0.964 | 1.279 [0.773, 2.800], *p*=0.488 |
| Maximum LFC | 0.766 [0.696, 0.856], ***p*<0.001** | 0.716 [0.631, 0.831], ***p*<0.001** | 0.765 [0.656, 0.927], ***p*<0.001** |
| Dataset Nonlinearity | 2.741 [1.470, 5.472], ***p*=0.002** | 4.850 [2.625, 9.575], ***p*<0.001** | 4.793 [2.428, 10.275], ***p*<0.001** |

**Multivariable Model Summary:**

| *Variable* | *XGB* | *RF* | *NN* |
| --- | --- | --- | --- |
| Intercept | 901.435 [726.034-1,128.885] | 539.432 [313.071-963.633] | 716.840 [451.136-1,191.176] |
| Minority Class Proportion | -- | 0.989 [0.976-1.002], *p*=0.086 | -- |
| Separability | 0.248 [0.187-0.329], ***p*<0.001** | -- | 0.499 [0.292-0.843], ***p*=0.011** |
| Number of Differentially Expressed Genes | -- | -- | -- |
| Average Read Count | -- | -- | -- |
| Dispersion | 1.381 [1.000-1.924], *p*=0.057 | 1.606 [0.988-2.628], *p*=0.055 | 2.325 [1.304-4.225], ***p*=0.008** |
| Correlation | -- | -- | -- |
| Median LFC | 0.563 [0.476-0.673], ***p*<0.001** | -- | -- |
| Minimum LFC | 1.655 [1.266-2.210], ***p*<0.001** | -- | -- |
| Maximum LFC | -- | 0.723 [0.663-0.790], ***p*<0.001** | 0.748 [0.665-0.854], ***p*<0.001** |
| Dataset Nonlinearity | -- | 3.307 [2.105-5.269], ***p*<0.001** | 2.436 [1.335-4.496], ***p*=0.007** |
| **Metrics** |  |  |  |
| Adj. Psuedo-R^2^ | 0.841 | 0.793 | 0.687 |

**Supplementary References:**

1. Heckerman D. A Tutorial on Learning With Bayesian Networks [Internet]. arXiv.org. 2020 [cited 2025 Aug 14]. Available from: https://arxiv.org/abs/2002.00269

2. Kaur D, Sobiesk M, Patil S, Liu J, Bhagat P, Gupta A, et al. Application of Bayesian networks to generate synthetic health data. Journal of the American Medical Informatics Association: JAMIA [Internet]. 2021 Mar 18 [cited 2022 Nov 16];28(4):801–11. Available from: <https://pubmed.ncbi.nlm.nih.gov/33367620/>

3. Glover F. Future paths for integer programming and links to artificial intelligence. Computers & Operations Research [Internet]. 1986 Jan [cited 2019 Nov 7];13(5):533–49. Available from: http://leeds-faculty.colorado.edu/glover/fred%20pubs/174%20-%20Future%20Paths%20for%20Integer%20Programming%20TS.pdf

4. Song W, Qin Z, Hu X, Han H, Li A, Zhou X, et al. Using Bayesian networks with Tabu-search algorithm to explore risk factors for hyperhomocysteinemia. Scientific Reports [Internet]. 2023 Jan 28;13(1):1610. Available from: https://www.nature.com/articles/s41598-023-28123-z

5. bnlearn - Bayesian network structure learning [Internet]. www.bnlearn.com. Available from: https://www.bnlearn.com/

6. Bates DM, Watts DG. Nonlinear Regression Analysis and Its Applications. Hoboken, New Jersey. Wiley-Interscience; 2007.

7. Kutner M, Nachtsheim C, Neter J. Applied Linear Regression Models. London. Mcgraw-Hill Education - Europe; 2004.

8. Bi R, Liu P. Sample size calculation while controlling false discovery rate for differential expression analysis with RNA-sequencing experiments. BMC Bioinformatics. 2016 Mar 31;17(1).

9. Jeon H, Xie J, Jeon Y, Kyeong Joo Jung, Gupta A, Chang W, et al. Statistical Power Analysis for Designing Bulk, Single-Cell, and Spatial Transcriptomics Experiments: Review, Tutorial, and Perspectives. Biomolecules [Internet]. 2023 Jan 24 [cited 2023 May 1];13(2):221–1. Available from: <https://www.ncbi.nlm.nih.gov/pmc/articles/PMC9952882/>

10. Poplawski A., Binder H. Feasibility of sample size calculation for RNA-seq studies. Brief. Bioinform. 2018;19:713–720. doi: 10.1093/bib/bbw144.
